# Comparative analysis of patient-derived organoids and patient-derived xenografts as avatar models for predicting response to anti-cancer therapy

**DOI:** 10.1101/2025.08.10.25333051

**Authors:** Joan Miguel Romero, Jamie Magrill, Nikita Kalashnikov, Yi Zhen Luo, Owen J. Chen, Sandrine Busque, Rong Ma, Aline Atallah, Anna-Maria Lazaratos, Daniel Mendelson, Liam Wilson, Shriya Deshmukh, Tarek Taifour, Mark Sorin, Isabella Arthur, Hellen Kuasne, Jeremy Y. Levett, Yifan Wang, Thomas Seufferlein, Alexander Kleger, Johann Gout, Alica K. Beutel, James Brugarolas, Zachry S. Poshusta, Tara L. Hogenson, Martin E. Fernandez-Zapico, Akinobu Hamada, Shigehiro Yagishita, Anthony Nichols, John W. Barrett, Federica Papaccio, Josefa Castillo, Masahiro Inoue, Thierry Massfelder, Hervé Lang, Veronique Lindner, Jonas Nilsson, Zahra Dantes, Gabrielle A. Wells, Sung Han Kim, Michael M. Ittmann, Hugo Villanueva, Seth P. Lerner, Andrew G. Sikora, Charles Theillet, Daniel Q. Huang, Dong-Anh Khuong-Quang, Jonathan C. Yeung, Peter M. Siegel, Ian R. Watson, George Zogopoulos, April A. N. Rose, Matthew Dankner

## Abstract

Patient-derived xenografts (PDX) and organoids (PDO) are widely used to model cancer and predict treatment response in matched patients. However, their predictive accuracy has not been systematically studied nor compared. We conducted a systematic review and meta-analysis of studies using PDX or PDO from solid tumors treated with identical anti-cancer agents as the matched patient, identifying 411 patient-model pairs (267 PDX, 144 PDO). Overall concordance in treatment response between patients and matched models was 70%, with no significant differences between PDX and PDO. Sensitivity, specificity, positive and negative predictive value were also comparable. Patients whose matched PDO responded to therapy had prolonged progression-free survival. For PDX, this association held only when analyses were restricted to patient-model pairs with low risk of bias after applying a bias assessment metric. Together, these findings suggest that PDO perform similarly to PDX in predicting matched-patient response, while potentially offering lower financial and ethical burdens.

**Graphical abstract:** 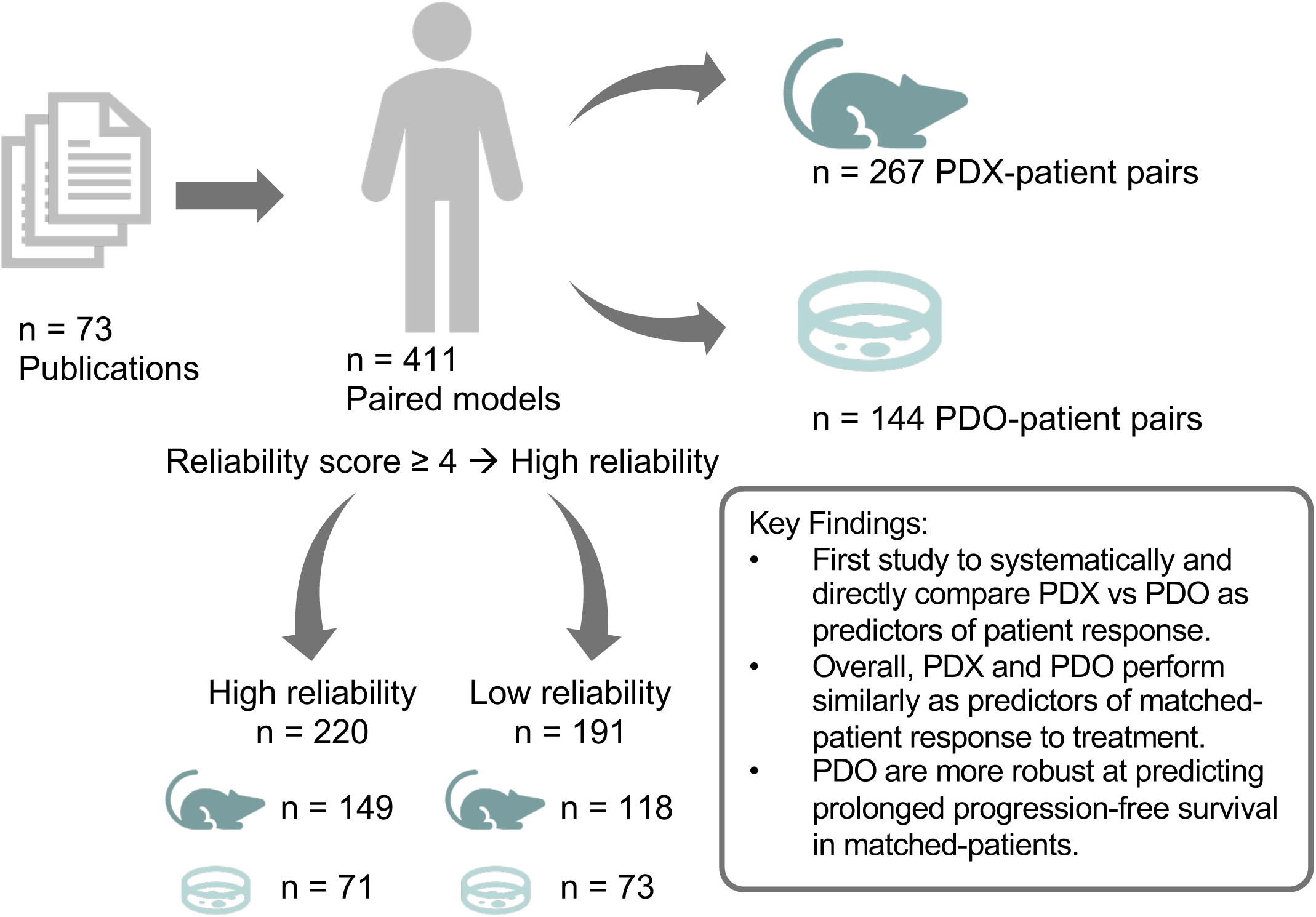

## INTRODUCTION

Patient avatar approaches to precision oncology have been extensively used in preclinical research but are rarely integrated into clinical practice due to limited evidence of clinical utility and challenges in establishing and testing these models efficiently^1–3^. Nonetheless, they remain central to translational cancer research and serve as foundational tools for the development of clinical trials^4^.

The two most widely studied avatar systems are patient-derived xenografts (PDX) and patient-derived organoids (PDO)^2,3^. PDX models involve implanting living human tumor tissue into immunocompromised hosts—typically mice—where the tumor retains much of its histological architecture and cellular heterogeneity while interacting with host stromal components^5–7^. PDX models offer important insights into tumor biology, drug response, and resistance mechanisms^8^. However, they are limited by the absence of an adaptive immune system and components of the microenvironment, while undergoing mouse-specific evolution^7,9^. They are also costly, time-consuming, and ethically complex due to the extensive use of animals.

PDO models, in contrast, involve culturing patient tumor cells as three-dimensional organoids *in vitro* using defined culture media supported by growth factors and extracellular matrix components^3^. While lacking the vascular and stromal complexity of *in vivo* systems, PDO can recapitulate key architectural and molecular features of the original tumor^3^. They offer a more cost-effective and scalable approach, are amenable to high-throughput drug screening, and avoid the ethical challenges of animal use^10,11^. However, the *in vitro* nature of PDO prevents them from being able to model tumor-stroma and immune cell interactions^11^. This limitation precludes PDO models from being used to study both immunotherapeutics and anti-angiogenic agents. Moreover, growth media additives required for PDO propagation may interfere with assessment of drug response^12^.

Both PDX and PDO models have been used to predict patient-specific treatment responses in relatively small, single-center studies^4^. However, no prior analysis has systematically assessed or directly compared their predictive performance. To address this, we conducted a systematic review and meta-analysis of matched patient-PDX/PDO treatment response pairs across solid cancers. This study offers key strategic insights for translational researchers, industry, and policy makers aiming to efficiently establish clinically meaningful patient-derived models. It also underscores ethical considerations, particularly regarding the widespread use of animals in PDX research, and informs future efforts to balance predictive accuracy, scalability, and feasibility in translational precision oncology.

## METHODS

### Search Strategy

A comprehensive literature search was conducted in MEDLINE and EMBASE for the period from January 1, 2002, to December 10, 2022, yielding 9,123 and 21,114 results, respectively (**Appendix 1**). Deduplication was performed using the *synthesisr* R package (v0.3.0) with optimal string alignment (restricted Damerau-Levenshtein distance, threshold = 2). Published conference abstracts were included. Additional relevant publications or data known to the authors outside of the database search were also considered. For abstracts with insufficient information to warrant inclusion without the accompanying full manuscript, corresponding authors were contacted for additional data. If no response was received, a follow-up email was sent at least 14 days later. If the required data could not be obtained, the abstract was excluded. The review protocol was prospectively registered with PROSPERO (ID: CRD42023387229) and conducted in accordance with the Preferred Reporting Items for Systematic Reviews and Meta-Analyses (PRISMA) guidelines^13^.

### Inclusion and exclusion criteria

Inclusion criteria were as follows: PDX and/or PDO models derived from human solid tumors treated with systemic therapies with response data reported, with matched-patient response data to the same agent(s). Tumors derived from both primary and metastatic sites were included. Studies including the use of any systemic anti-cancer therapeutics approved by the United States Food and Drug Administration for human use were included. Any studies of patients examining only PDX and/or PDO models without accompanying matched patient response data were excluded. Liquid tumors, non-murine PDX models, models derived through an intermediate step rather than directly from the patient (i.e., organoids derived from a PDX, or vice versa) or studies of patients who received local therapies, including radiation, as part of the treatment line of interest, were excluded. Patient-model pairs treated with hormone therapies or immunotherapies were excluded due to limited sample sizes. Additionally, the mechanisms of action of immunotherapies are predominantly tumor-extrinsic by involving the tumor microenvironment, which limits the feasibility of comparing their effects across PDX and PDO models.

### Abstract screening

All abstracts were screened in duplicate by independent reviewers (JMR, JM, NK, ML, OC, AA, SD, TT, MS, MD) using *Covidence*. References were managed using *EndNote X9*. Conflicts were resolved through discussion among the three chief reviewers (JMR, JM, MD). Abstracts were assessed for the presence of treatment response data from PDX and/or PDO models with corresponding matched-patient response data. Exclusion criteria at the abstract stage included: (1) no reported treatment administration to PDX/PDO models, (2) absence of text suggesting the possibility of matched patient response information in the full manuscript, (3) conference abstracts duplicating data later published in full manuscripts, and (4) review articles.

### Data extraction

Data extraction was conducted by independent reviewers from the author team (JMR, JM, NK, ML, OC, AA, SD, TT, MS, MD), with each full-text publication reviewed by two individuals. Discrepancies were resolved through discussion; if consensus could not be reached, one of the three chief reviewers (JMR, JM, MD) made the final determination.

PDX and PDO treatment response was recorded as defined by the original authors. To account for heterogeneity of response reporting practices, a risk of bias assessment was applied to separate patient-model pairs based on reporting quality (discussed further below). Patient treatment response was assessed using RECIST v1.1 criteria when reported; otherwise, author-determined criteria were used. Progression-free survival (PFS) was used as a secondary clinical endpoint when available.

All patient-model pairs included models treated with a single specified dose. When multiple doses were tested for an individual PDX model, the closest murine equivalent of the clinical dose was selected using the conversion method by Nair and Jacob, and only that pair was included in downstream analysis^14^. This adjustment was only applicable in n=13 instances. Both subcutaneous and orthotopic PDX models were eligible; the only orthotopically implanted PDX models eligible for inclusion were n=4 breast cancer primaries implanted into the mouse mammary fat pad. For studies reporting only aggregate data, corresponding authors were contacted to request individual-level data. If no response was received, a follow-up email was sent at least 14 days later.

### Quality (risk of bias) assessment

To assess the methodological quality of individual studies included in the meta-analysis, we adapted a previously described tool derived from the Newcastle-Ottawa scale^15,16^. The tool included 6 items, covering the selection and representativeness of cases, the ascertainment of treatments, origins and outcomes, patient follow-up and reporting, and technical replicates of PDX and PDO, with each item scored one point if the information was specifically reported. All included studies and patient-model pairs were assessed for risk of bias by a single reviewer (JM, AL, LW, or DM). In cases of uncertainty, a final decision was made by one of the three chief reviewers (JMR, JM, MD). Patient-model pairs or studies were classified as high reliability (low risk of bias) if they met at least 4 of the 6 criteria (score ≥4), and as low reliability (high risk of bias) if they met 3 or fewer criteria (score ≤3).

The detailed quality assessment criteria used were as follows:

1. *Selection Bias: Did the paper include treatment responses from all PDX or PDO-matched patient pairs from the modeling project?*
2. *Validation: Was the PDX or PDO successfully validated to originate from matching human tissue with a DNA, RNA, or protein-based assay, or by histology?*
3. *Patient Outcome: Was patient response determined by RECIST criteria, and was the PDX or PDO response data obtained objectively?*
4. *Follow-Up Length: Was patient follow-up sufficiently long enough for meaningful survival times to be evaluated (a survival time of at least 6 months, or earlier in the event of early death)?*
5. *Sufficient Reporting: Was the PDX or PDO-matched patient pair described with sufficient detail? This criterion is comprised of 4 sub-categories, each of which needed to be reported for the criterion to be scored:*

- *Route of administration for the model is confirmed to be the same as for the patient.*
- *Dosing regimen is described for the patient.*
- *Dosing regimen is described for the PDX or PDO.*
- *Previous lines of systemic treatment are described for the patient.*
6. *Replicates: Were there at least n = 3 replicates for the PDX or PDO?*

### Statistical analyses

Survival analyses were performed using the Kaplan-Meier estimator to predict survival probabilities and log-rank test for statistical comparisons, using the survival (v3.7.0) package. One-tailed P-values for survival curves (Figure 4, Supplementary Figures 5-7) were calculated as follows: when curves followed the alternative hypothesis in the expected direction (patient had longer PFS when matched model did respond), the one-tailed P-value equaled the two-tailed P-value divided by 2; when the curves followed the alternative hypothesis in the unexpected direction (patient had longer PFS when matched model did not respond), the one-tailed P-value equaled 1 minus half of the two-tailed P-value. Curves were determined to follow the alternative hypothesis (patients whose model responded predicted survival) when the latter group had higher median progression-free survival than models that did not respond. Hazard ratios and statistics were calculated using a multivariate Cox proportional hazard model from the survival (v3.7.0) package and plotted using the forestmodel (v.0.6.2) package. For survival analyses, the following variables were used in the multivariate Cox Proportional Hazard model: response, cancer type, and treatment type. Cancer and treatment types with at least 3 patient-model pairs per variable were included for multivariate analysis. After applying this filtering, some combinations had fewer than three patient-model pairs due to overlap between the excluded cancer and/or drug types. These were retained to allow for comparison of as many variables possible. Specificity, sensitivity, positive predictive value (PPV) and negative predictive value (NPV) were calculated using the epiR (v2.0.74) package and confirmed by manual calculation. Fisher’s exact test was used to compare frequencies. False discovery rate (FDR) was calculated using the Benjamini-Hochberg procedure and used to correct for multiple testing where indicated. All statistical analysis and graphic representation of data was performed in the R statistical programming language (v4.2.1), and respective packages from the CRAN repositories.

## RESULTS

### Cohort demographics

Following screening and data extraction from 21,565 abstracts, a total of 73 publications containing 411 patient-model pairs were included in the final analysis (**Figure 1**, **Appendix 2**). Of the 411 patient-model pairs, 267 (65%) were PDX, and 144 (35%) were PDO (**Table 1**). Across all model-patient pairs, the majority of PDX came from North America (160/267, 60%), while the majority of PDO were modeled in Asia (114/144, 79%, **Table 1**). Most PDX and PDO were from patients with metastatic disease, with no difference between models (136/182 PDX, 74.7%; 96/113 PDO, 85.0%, not significant, ns).

**Figure 1.**
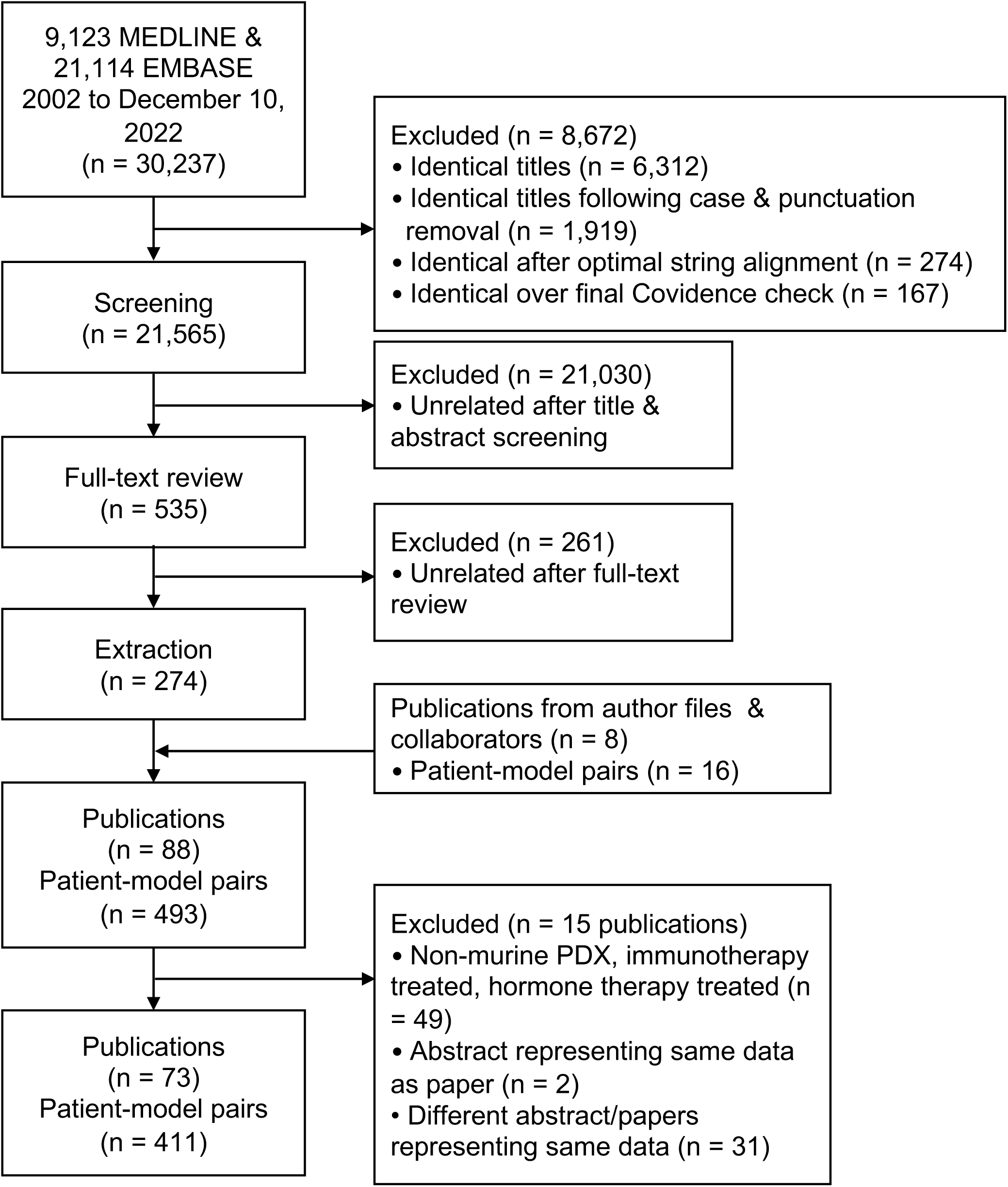
Consort diagram. Following screening, full-text review, and extraction, 411 patient-model pairs were extracted from 73 publications.

**Table 1.** Cohort Demographics of PDX and PDO patient-model pairs. Demographic variables compared between PDX and PDO patient-model pairs. For cancer type, top six cancer types based on total N are shown. Percentages of total for category shown in brackets. P-values were calculated using Fisher’s exact test and were corrected to adjusted-P values calculated using the Benjamini-Hochberg procedure. Abbreviations: PDX; patient-derived xenograft, PDO; patient-derived organoid, NSCLC; non-small cell lung cancer, NS; not significant.

|  | Total | PDX | PDO | P-adj Value |
| --- | --- | --- | --- | --- |
| All | 411 | 267 (65%) | 144 (35%) | - |
| **Continent** |  |  |  |  |
| North America | 172 | 160 (93%) | 12 (7%) | < 0.001 |
| Asia | 197 | 83 (42%) | 114 (58%) | < 0.001 |
| Europe | 40 | 24 (60%) | 16 (40%) | NS |
| Australia | 2 | 0 (0%) | 2 (100%) | NS |
| **Sex** |  |  |  |  |
| Male | 159 | 88 (55%) | 71 (45%) | NS |
| Female | 130 | 87 (67%) | 43 (33%) | - |
| **Age, mean (min-max)** |  |  |  |  |
|  | 54.34 (1-83) | 52.56 (1-79) | 57.35 (18-83) | < 0.05 |
| **Stage** |  |  |  |  |
| Metastatic | 232 | 136 (59%) | 96 (41%) | NS |
| Non-metastatic | 63 | 46 (73%) | 17 (27%) | - |
| **Cancer** |  |  |  |  |
| Colorectal | 102 | 20 (20%) | 82 (80%) | < 0.001 |
| Sarcoma | 26 | 26 (100%) | 0 (0%) | < 0.001 |
| NSCLC | 49 | 28 (57%) | 21 (43%) | NS |
| Ovarian | 77 | 75 (97%) | 2 (3%) | < 0.001 |
| Breast | 31 | 28 (90%) | 3 (10%) | < 0.01 |
| Pancreatic | 25 | 20 (80%) | 5 (20%) | NS |
| Other | 101 | 70 (69%) | 31 (31%) | NS |
| **Treatment** |  |  |  |  |
| Chemotherapy | 244 | 147 (60%) | 97 (40%) | < 0.05 |
| Targeted therapy | 122 | 87 (71%) | 35 (29%) | NS |
| Combination therapy | 45 | 33 (73%) | 12 (27%) | NS |

The most modelled cancer types were colorectal, followed by ovarian, NSCLC, and breast cancer (**Figure 2A**). Colorectal cancer was predominantly modeled using PDO (82/102, 80%, P < 0.001, **Table 1**, **Figure 2A**) while breast and ovarian cancers were primarily modeled using PDX (Breast: 28/31, 90%, P = 0.004; Ovarian: 75/77, 97%, P < 0.001 **Table 1**, **Figure 2A**). NSCLC was similarly represented in both PDX and PDO (28/49 PDX, 57% vs. 21/49 PDO, 43%, not significant [ns], **Table 1**, **Figure 2A**). The majority of PDX and PDO were treated with cytotoxic chemotherapy alone, with more PDO treated with chemotherapy alone compared to PDX (147/267 PDX, 55% vs. 97/144 PDO, 67%, P < 0.05, **Table 1**). This was followed by targeted therapies, with no difference between models (87/267 PDX, 33%; 35/144 PDO, 24%, ns, **Table 1**). 45 patient-model pairs were treated with a combination of chemotherapy and targeted therapy (33/267 PDX, 12%; 12/144 PDO, 8%, ns, **Table 1).**

**Figure 2.**
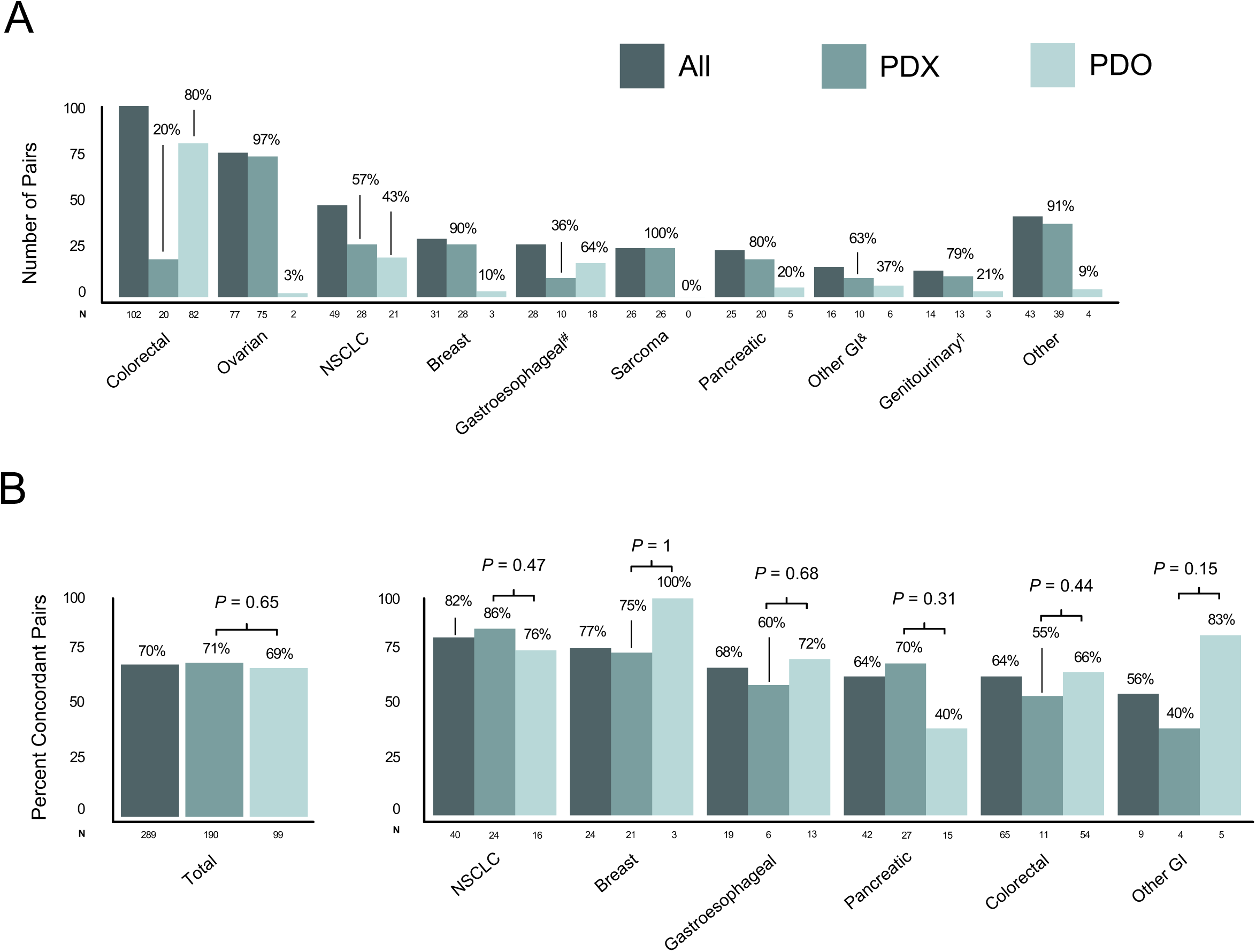
Demographic overview of cohort and concordance. A) Distribution of PDX and PDO patient-model pairs across grouped cancer types. ^#^Gastroesophageal PDX = 8 gastroesophageal, 1 esophageal, 1 gastric; PDO = 15 gastric, 2 esophageal, 1 gastroesophageal. ^&^Other GI PDX = 3 cholangiocarcinoma, 3 hepatocellular, 2 duodenal, 2 anal; PDO = 3 gastroenteropancreatic neuroendocrine, 1 cholangiocarcinoma, 1 hepatocellular, 1 small bowel. ^†^Genitourinary PDX = 4 renal, 3 urothelial, 2 prostate, 1 bladder, 1 testicular; PDO = 2 neuroendocrine prostate, 1 testicular. B) Percent of patient-model pairs with concordant responses compared to matched patients for all (left) and across cancers with at least 3 pairs per type (right). P-values calculated with Fisher’s exact test. Abbreviations: PDX; patient-derived xenograft, PDO; patient-derived organoid, NSCLC; non-small cell lung cancer, GI; gastrointestinal.

We then compared patient response to treatment between models across all 411 patient-model pairs. Patients from whom PDX were generated were more likely to have a clinical response to treatment, defined as complete or partial response (CR or PR), compared to those whose cancer was modeled using PDO (123/267 PDX, 46% vs. 48/144 PDO, 33%, P = 0.016, **Supplementary Figure 1A, B**). We next assessed treatment response rate of models across individual cancer and treatment types. Cancers including breast, gastroesophageal, pancreatic, and NSCLC had similar response rates across both PDX and PDO (**Supplementary Table 1, Supplementary Figure 1C**). Colorectal cancers (PDX: 16/20, 80%; PDO: 41/82, 50%, P = 0.02) showed differences in response to treatment between PDX and PDO (**Supplementary Table 1, Supplementary Figure 1C**). PDX had higher response rates when treated with chemotherapy (PDX: 100/147, 68%; PDO: 49/97, 51%, P = 0.007) or combination (PDX: 28/33, 85%; PDO: 6/12, 50%, P = 0.04) regimens containing chemotherapy with targeted therapies, while PDO had higher response rates when treated with targeted therapies compared to PDX (PDX: 40/87, 46%; PDO: 24/35, 69%, P = 0.03) (**Supplementary Figure 1D**).

### PDX and PDO exhibit similar predictive performance metrics

We next assessed the ability of PDX and PDO models to predict patient outcomes in the context of a given treatment. In the cohort at large, PDX and PDO had similar rates of concordance in responsiveness to treatment when compared to matched patients (190/267 PDX, 71% vs. 99/144 PDO, 69%, ns, **Table 2**, **Figure 2B**). There were no differences in concordance between PDX and PDO in any individual cancer type and treatment type, nor by sex, geography, or disease stage (**Table 2**, **Figure 2B**, **Supplementary Figure 1E and F**). Similarly, PDX and PDO demonstrated no significant differences in sensitivity or specificity (87% vs. 85%, ns; 58% vs. 60%, ns), nor PPV or NPV (64% vs. 52%, ns; 84% vs. 89%, ns, **Figure 3A**). Across cancer histologies, we did not observe any individual cancer exhibiting significant differences in sensitivity, specificity, PPV and NPV in PDX versus PDO (**Supplementary Figure 2**).

**Figure 3.**
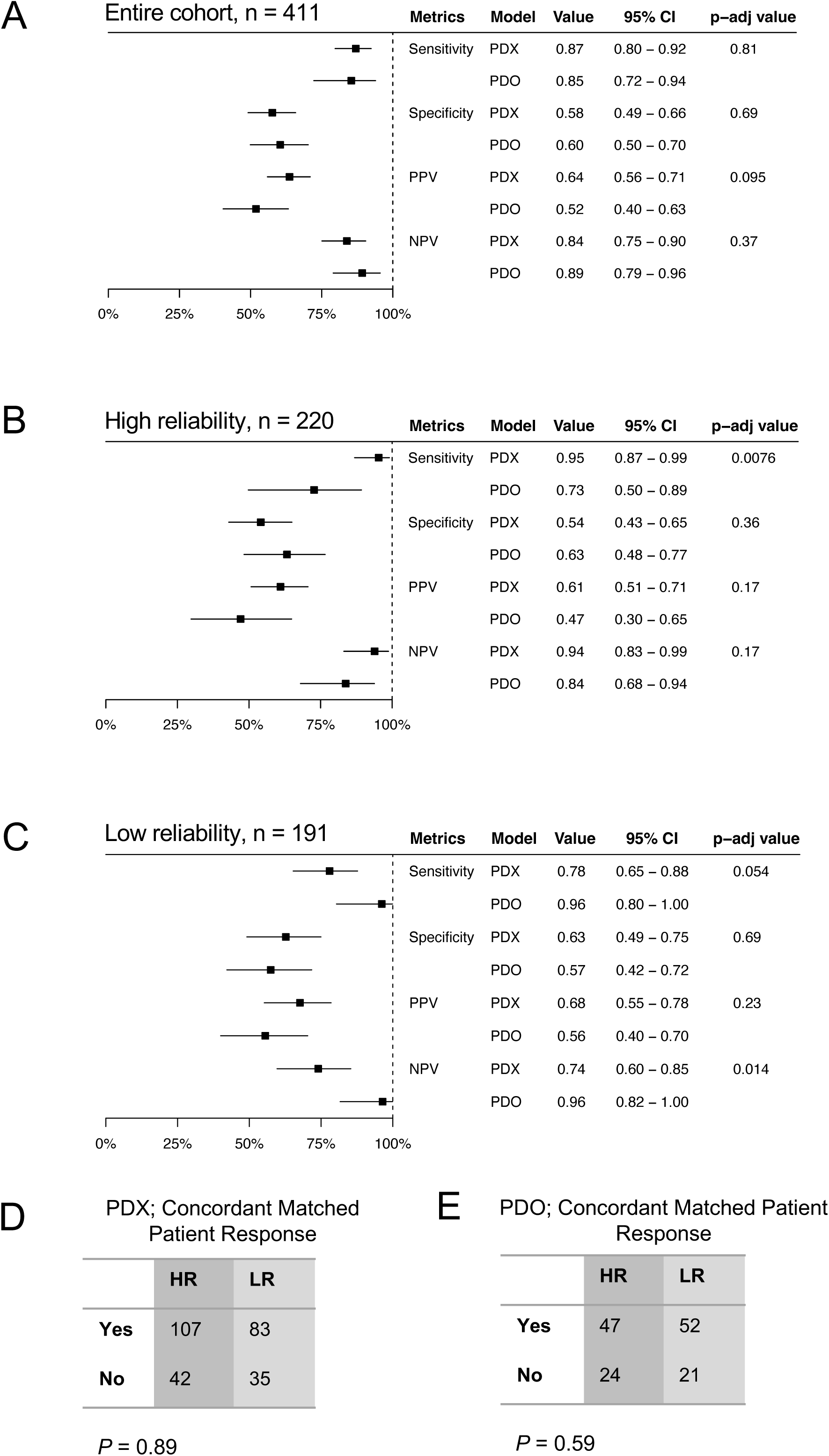
Predictive statistics comparing PDX and PDO. A) Sensitivity, specificity, PPV and NPV between PDX and PDO. Point estimates, 95% CI, and adjusted P-values are shown. B) Predictive statistics between PDX and PDO, for high reliability (HR) and C) low reliability (LR) patient-model pairs. P-values were calculated with Fisher’s exact test and corrected with the Benjamini-Hochberg procedure to calculate adjusted P-values. D) Contingency tables highlighting concordance rates between high HR and E) LR pairs. P-values calculated with Fisher’s exact test. Abbreviations: PDX; patient-derived xenograft, PDO; patient-derived organoid, PPV; positive predictive value, NPV; negative predictive value, CI; confidence interval.

**Table 2.** Concordance segregated by clinical variables. Comparison of concordance across clinical variables, for the total cohort, PDX, and PDO. For cancer type, top six cancer types based on total N are shown. P-values were calculated using Fisher’s exact test and were corrected to adjusted-P values using the Benjamini-Hochberg procedure. Abbreviations: PDX; patient-derived xenograft, PDO; patient-derived organoid, NSCLC; non-small cell lung cancer, NS; not significant.

|  | Total |  |  |  | PDX |  |  |  | PDO |  |  |  | PDX vs PDO |
| --- | --- | --- | --- | --- | --- | --- | --- | --- | --- | --- | --- | --- | --- |
|  | Total | Concordant | Non-concordant | P-adj Value | Total | Concordant | Non-concordant | P-adj Value | Total | Concordant | Non-concordant | P-adj Value | P-adj Value |
| All | 411 | 289 (70%) | 122 (30%) | - | 267 | 190 (71%) | 77 (29%) | - | 144 | 99 (69%) | 45 (31%) | - | NS |
| Continent |  |  |  |  |  |  |  |  |  |  |  |  |  |
| North America | 172 | 121 (70%) | 51 (30%) | NS | 160 | 114 (71%) | 46 (29%) | NS | 12 | 7 (58%) | 5 (42%) | NS | NS |
| Asia | 197 | 141 (72%) | 56 (28%) | NS | 83 | 55 (66%) | 28 (34%) | NS | 114 | 86 (75%) | 28 (25%) | < 0.05 | NS |
| Europe | 40 | 26 (65%) | 14 (35%) | NS | 24 | 21 (88%) | 3 (12%) | NS | 16 | 5 (31%) | 11 (69%) | < 0.05 | <0.01 |
| Australia | 2 | 1 (50%) | 1 (50%) | NS | - | - | - | - | 2 | 1 (50%) | 1 (50%) | NS | - |
| Sex |  |  |  |  |  |  |  |  |  |  |  |  |  |
| Male | 159 | 119 (75%) | 40 (25%) | NS | 88 | 68 (77%) | 20 (23 %) | NS | 71 | 51 (72%) | 20 (28%) | NS | NS |
| Female | 130 | 88 (68%) | 42 (32%) | - | 87 | 58 (67%) | 29 (33%) | - | 43 | 30 (70%) | 13 (30%) | - | - |
| Stage |  |  |  |  |  |  |  |  |  |  |  |  |  |
| Metastatic | 232 | 162 (70%) | 74 (30%) | NS | 136 | 96 (71%) | 40 (29%) | NS | 96 | 66 (69%) | 30 (31%) | NS | NS |
| Non-metastatic | 63 | 49 (78%) | 14 (22%) | - | 46 | 39 (85%) | 7 (15%) | - | 17 | 10 (59%) | 7 (41%) | - | - |
| Cancer |  |  |  |  |  |  |  |  |  |  |  |  |  |
| Colorectal | 102 | 65 (64%) | 37 (36%) | NS | 20 | 11 (55%) | 9 (45%) | NS | 82 | 54 (66%) | 28 (34%) | NS | NS |
| Sarcoma | 26 | 20 (77%) | 6 (23%) | NS | 26 | 20 (77%) | 6 (23%) | NS | - | - | - | - | - |
| NSCLC | 49 | 40 (82%) | 9 (18%) | NS | 28 | 24 (86%) | 4(14%) | NS | 21 | 16 (76%) | 5 (24%) | NS | NS |
| Ovarian | 77 | 49 (64%) | 28 (36%) | NS | 75 | 49 (65%) | 26 (35%) | NS | 2 | 0 (0%) | 2 (100%) | NS | NS |
| Breast | 31 | 24 (77%) | 7 (23%) | NS | 28 | 21 (75%) | 7 (25%) | NS | 3 | 3 (100%) | 0 (0%) | NS | NS |
| Pancreatic | 25 | 16 (64%) | 9 (36%) | NS | 20 | 14 (70%) | 6 (30%) | NS | 5 | 2 (40%) | 3 (60%) | NS | NS |
| Other | 101 | 75 (74%) | 26 (26%) | NS | 70 | 51 (73%) | 19 (27%) | NS | 31 | 24 (77%) | 7 (23%) | NS | NS |
| Treatment |  |  |  |  |  |  |  |  |  |  |  |  |  |
| Chemotherapy | 244 | 172 (70%) | 72 (30%) | NS | 147 | 106 (72%) | 41 (28%) | NS | 97 | 66 (68%) | 31 (32%) | NS | NS |
| Targeted therapy | 122 | 79 (65 %) | 43 (35%) | NS | 87 | 57 (66%) | 30 (34%) | NS | 35 | 22 (63%) | 13 (37%) | NS | NS |
| Combination therapy | 45 | 38 (84%) | 7 (16%) | NS | 33 | (82%) | 6 (18%) | NS | 12 | 11 (92%) | 1 (8%) | NS | NS |

### The application of quality assessment criteria elucidates differences in the predictive ability of PDX and PDO

Given differences in applications, cost, and throughput, we hypothesized that intrinsic biases exist for PDX and PDO models which, if controlled for, could clarify our understanding of their utility as patient avatars in predicting response to treatment. We adapted a 6-criterion scoring system from the Newcastle-Ottawa scale to evaluate reliability and risk of bias across the 411 individual patient-model pairs. Using this scoring system, we classified patient–model pairs as high reliability (HR) or low reliability (LR) based on a cutoff of ≥4 criteria met, resulting in 220 HR and 191 LR patient-model pairs (**Supplementary Figure 3**).

When comparing baseline characteristics between HR and LR pairs, colorectal, pancreatic, gastroesophageal, other gastrointestinal, and other cancer PDX were more likely to be HR compared to the cohort at large (P < 0.05; **Supplementary Table 2, Supplementary Figure 4**). While there was no significant difference between PDX and PDO in their respective proportions of HR and LR patient-model pairs (HR: 149/267 (55%), PDX vs. 71/144 (49%), PDO, ns), we found that studies using PDO had higher scores in meeting the criteria for ascertainment (P < 0.001), and follow up time (P < 0.001), while PDX-containing studies were more likely to have more detailed treatment reporting (P = 0.001), and technical replicates (P < 0.001) (**Supplementary Figure 3**). This suggests that overall, studies using PDX were more narrowly focused with more abundant patient treatment details and replicates, while PDO-containing studies were of higher throughput with less focus on individual patient-model pairs.

We next compared performance characteristics between PDX and PDO across HR and LR patient-model pairs. As was the case in the cohort at large, there were no differences in concordance between PDX and PDO when analyses were restricted to HR or LR pairs (HR: 107/149 (72%), PDX vs. 47/71 (66%), PDO, ns; LR: 83/118 (70%), PDX vs. 52/73 (71%), PDO, ns; **Supplementary Tables 3 and 4**). When analyses were restricted to HR pairs, PDX were more sensitive as compared to PDO (95% versus 73%, P = 0.008, **Figure 3B**), and when analyses were restricted to LR pairs, PDO had higher NPV as compared to PDX (96% versus 74%, P = 0.014, **Figure 3C**). No differences were observed in concordance rates between HR and LR patient-model pairs in either PDX or PDO (**Figure 3D, E**). Similarly, no difference in other demographic variables and concordance were observed between PDX and PDO patient-model pairs in HR and LR datasets (**Supplementary Tables 3 and 4**).

### Response of PDO models to treatment robustly predicts prolonged progression-free survival in matched patients

Given that response rate is only one measure of patient outcome, we sought to test the hypothesis that responsiveness of a model system is associated with prolonged PFS in the matched patient. Indeed, among 176 patient-model pairs with available patient PFS data, response to treatment in the model system was associated with prolonged PFS (median PFS; mPFS [95% confidence interval (CI)], responders: 10.4 [8.8-13.7] months vs. non-responders: 5.0 [4.4-9.1], months, P = 0.02) (**Figure 4A**). This finding remained in multivariate analyses when controlling for cancer and treatment type (Hazard Ratio, [95% CI]: 0.39 [0.26-0.58], P < 0.001, **Figure 4A**).

**Figure 4.**
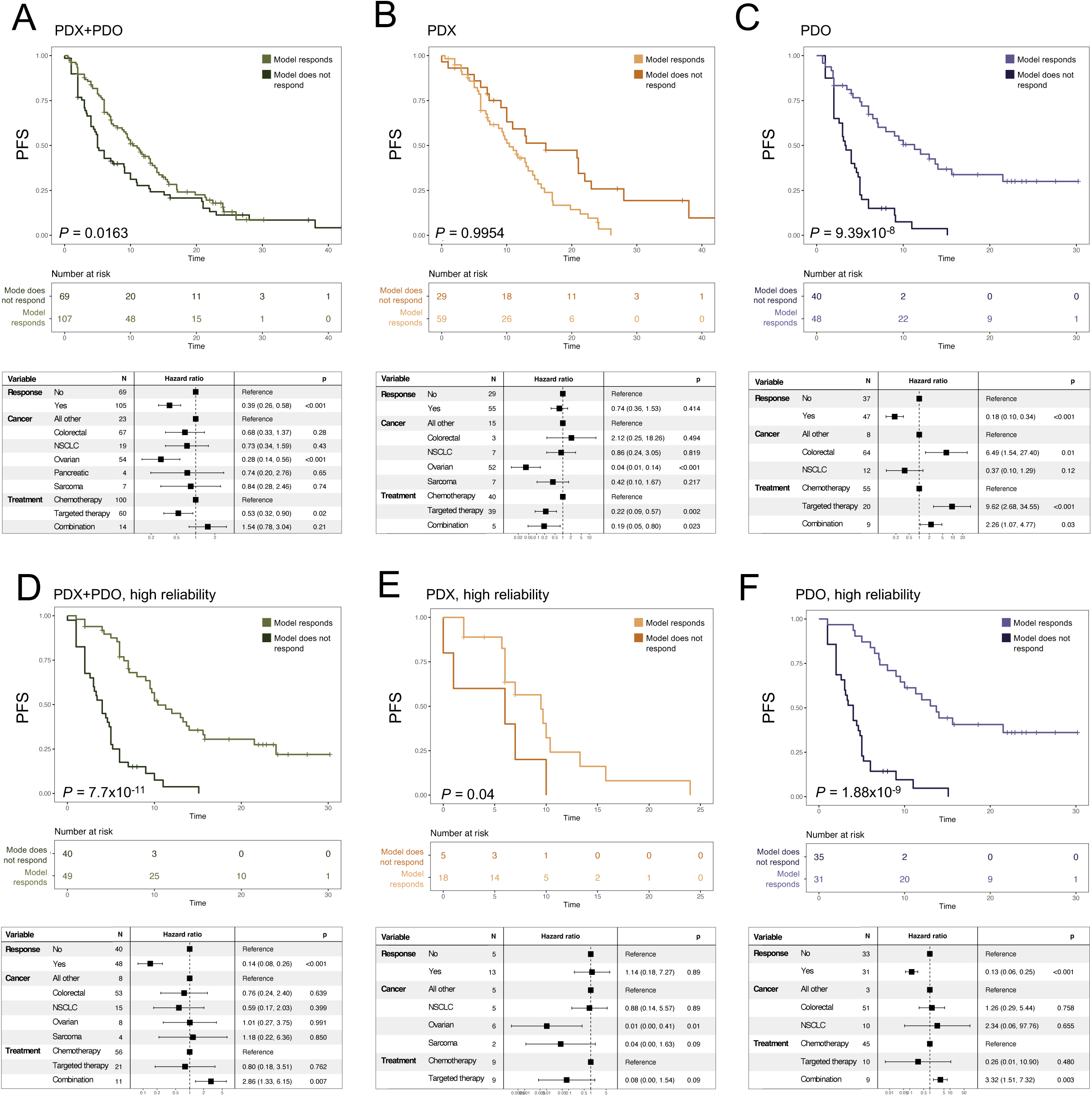
Responsiveness in model is associated with prolonged progression-free survival in matched patient. Comparison of PFS in patients from whom associated model did or did not respond to treatment across A) entire cohort, B) PDX and C) PDO. Comparison of PFS in patients from high reliability patient-model pairs and from whom associated model did or did not respond to treatment across D) entire cohort, E) PDX, and F) PDO. One sided P-values calculated using log-rank test (Methods). Below each Kaplan-Meier curve are multivariate Cox proportional hazard models assessing the association between variables and patient PFS after removing categories with insufficient sample size (Methods). Abbreviations: PDX; patient-derived xenograft, PDO; patient-derived organoid, NSCLC; non-small cell lung cancer, PFS; progression-free survival.

When analyses were restricted by model type, PDO patient-model pairs (responders: mPFS 11.3 [7.0-21.5] months vs. non-responders: mPFS 3.4 [2.6-5.0] months, P < 0.001), but not PDX patient-model pairs (responders: mPFS 10.4 [7.4-14.0] months vs. non-responders: mPFS 16.0 [10.0-28.0] months, ns) responding to treatment were associated with prolonged patient PFS in both univariate (log-rank) and multivariate analyses (PDX: 0.74 [0.36-1.53], ns; PDO: 0.18 [0.10-0.34], P < 0.001, **Figure 4B, C**).

When analyses were restricted based on reliability score, we observed strengthening of the signal in both univariate (responders: 10.4 [9.5-15.8] months vs. non-responders: 4.0 [3.0-5.0] months, P < 0.001) and multivariate analyses (0.14 [0.08-0.26], P < 0.001) in the cohort at large in HR pairs (**Figure 4D**). For PDX patient-model pairs, limiting analyses to only HR pairs resulted in prolonged PFS in patients from whom the matched model responded to treatment (responders: mPFS 9.5 [6.0-NA] months vs. non-responders: mPFS 6.0 [1.0-NA] months, P = 0.04, **Figure 4E**). However, this statistical significance did not persist in multivariate analyses (1.14 [0.18-7.27], ns, **Figure 4E**). When analyzing only HR PDO patient-model pairs, responders demonstrated prolonged PFS in both univariate (responders: mPFS 13.7 [9.5-NA] months vs. non-responders: mPFS 4.00 [3.0-5.0] months, P < 0.001) and multivariate analyses (0.13 [0.06-0.25], P < 0.001, **Figure 4F**).

When analyses were restricted to LR pairs only, there was no difference in PFS in univariate (responders: 11.0 [6-14.3] vs. non-responders: 16.0 [10-28.0], ns) or multivariate analyses (1.42 [0.70-2.85], ns) between patients whose models responded versus those whose model did not respond to treatment in the total cohort. (**Supplementary Figure 5A**). The same was observed when analyses were restricted to LR PDX for both univariate (responders: 11.7 [8.75-17.0] vs. non-responders 21.0 [13.0-NA], ns) and multivariate analyses (0.74 [0.31-1.79], ns, **Supplementary Figure 5B**). The association between response and prolonged PFS was also lost for LR PDO patient-model pairs in both univariate (responders: 10 [5.2-NA] vs. non-responders: 2.0 [2.0-NA], ns) and multivariate analyses (0.3 [0.07-1.37], ns, **Supplementary Figure 5C**).

Given the demonstrated value of applying quality assessment criteria to PDX and PDO models to improve their ability to predict prolonged PFS in responsive models, we next sought to determine which, if any, individual criteria, were most important in driving this statistical signal. We repeated our analyses for PDX-patient pairs based on each individual criteria, with only criteria 2 (patient-model matching validation) replicating the signal observed in the cohort of HR patient-model pairs in univariate analysis (response: 8.75 [6.0-15.8] vs. no response: 5.50 [1.0-NA], P = 0.03, **Supplementary Figure 6**). None of the individual criteria achieved statistical significance in multivariate analyses. Meanwhile, PDO that responded to treatment were associated with longer matched patient PFS across all evaluable quality assessment metrics in both univariate and multivariate analyses (P < 0.001) (**Supplementary Figure 7**). Taken together, these results suggest response to treatment in PDO models can reliably predict prolonged patient PFS to matched treatment, while PDX models can achieve this only when specific quality assessment metrics are met.

## DISCUSSION

Preclinical models used to predict patient response to anti-cancer therapies are important tools for informing clinical trial design and therapeutic development^2^. Among these, PDX and PDO are the most widely used patient avatar models for assessing treatment efficacy in matched patients^4^. While several small studies have demonstrated the predictive value of both models across various cancer types^17–22^, no study to date has systematically evaluated their performance or directly compared them in a head-to-head manner. Thus, the objectives of this study were to conduct a systematic review and meta-analysis of studies reporting treatment responses in PDX and PDO models with matched-patient outcomes, and to identify key methodological and reporting criteria necessary to ensure accurate and reproducible modeling of clinical response.

We found that PDX and PDO had similar predictive performance statistics, with both having concordance rates of approximately 70% and no differences in sensitivity, specificity, PPV nor NPV. Importantly, response to treatment in PDO, but not PDX, were able to predict prolonged PFS in matched patients in the cohort at large.

Differences in model performance emerged after applying a predefined quality assessment threshold to stratify patient-model pairs into high reliability (HR) and low reliability (LR) cohorts. When analyses were restricted to HR pairs, PDX demonstrated greater sensitivity than PDO models. Additionally, treatment response in both PDX and PDO models was associated with longer PFS in matched patients in univariate analyses.

While PDX models offer unique scientific advantages, such as preserving tumor histopathology and enabling investigation of tumor–microenvironment interactions ^23,24^, we found a lack of compelling evidence to support their superiority over PDO models in predicting matched-patient treatment response. Similarly, stratified analyses by cancer histology and treatment type did not reveal significant differences in predictive performance between PDX and PDO, although limited sample sizes in several subgroups may have constrained statistical power. Given these findings and considering the higher financial costs and ethical burdens associated with PDX models, we propose that PDO models may be generally preferred for the specific application of patient avatars in precision oncology. These conclusions align with recent initiatives by the U.S. National Institutes of Health aimed at reducing animal use in biomedical research^25^.

While this study represents a rigorous meta-analysis of the published literature on PDX and PDO models as avatar predictors of patient outcomes, several important limitations merit discussion. Most notably, treatment response assessments for both patient and model relied on author-defined criteria rather than a standardized methodology. This limitation reflects the heterogeneity of response classification systems across studies and the lack of consistency in reported raw data. To mitigate this, we incorporated PFS as a secondary clinical endpoint and applied a structured risk of bias assessment to account for variability in study quality and reporting.

As the patient avatar field continues to mature, the adoption of standardized, internationally recognized reporting criteria will be critical for improving reproducibility, enabling cross-study comparisons, and accelerating clinical translation of avatar-based precision oncology. Although reporting guidelines have recently been proposed for both PDX and PDO models, they have not yet been widely implemented^26,27^. In this context, we propose a 6-point reporting framework used in this study as a practical and informative checklist for researchers, particularly when publishing studies involving PDX models. This is supported by our finding that response to treatment only in HR, but not LR, PDX models was associated with prolonged patient PFS. Furthermore, inherent differences in pharmacokinetics and pharmacodynamics between humans, murine, and *in vitro* organoid systems present a significant challenge in comparing model and patient responses. In this study, we relied on author-reported dosing, as standardized dose equivalency data were rarely available. To partially address this limitation, we applied a published human-to-mouse dose conversion method in cases where multiple doses were used in PDX models^14^, selecting the dose most closely aligned with human dosing. However, this adjustment was only applicable in 13 instances, limiting its overall impact. These differences underscore the need for more consistent reporting and standardized dosing strategies in future preclinical trials.

Another limitation of this study is the lack of standardization in the generation of PDX and PDO models across studies. Given the diversity of techniques used to establish these models, we included all murine PDX, and all PDO models to maximize statistical power and assess potential differences in predictive performance. This included PDX models implanted both subcutaneously and orthotopically. However, only four orthotopic PDX models met inclusion criteria, all of which derived from breast cancer primary tumors that were implanted in the mouse mammary fat pad. Notably, no orthotopic models were derived from other primary tumor sites or implanted in anatomically relevant metastatic sites. This represents a significant limitation of the existing literature, as orthotopic implantation may more accurately recapitulate the tumor microenvironment and drug response of the matched patient. Furthermore, emerging species used for generating PDX, such as zebrafish, have gained increased attention in recent years; these species were not included in this analysis due to small sample size in the published literature compared to murine PDX models^28^.

Taken together, the results of this study suggest that while both PDX and PDO models predict patient response to anti-cancer therapies with similar overall accuracy, PDO models more robustly predict prolonged PFS in matched patients. Importantly, applying a standardized quality assessment framework improved the predictive performance of both PDX and PDO, underscoring the value of methodological rigor in avatar-based research. As the use of PDX, PDO, and emerging platforms including organ-on-chip technologies and orthotopically implanted PDX models continues to grow in patient-response prediction and drug discovery, future studies will need to incorporate larger sample sizes, standardized quality metrics, and robust reporting practices. These efforts will be essential to reassess and refine the role of these models in precision oncology.

## Data Availability

All data produced in the present study are available upon reasonable request to the authors

## Ethical Statement

The authors are accountable for all aspects of the work in ensuring that questions related to the accuracy or integrity of any part of the work are appropriately investigated and resolved. The study was conducted in accordance with the Declaration of Helsinki (as revised in 2013).

## SUPPLEMENTARY FIGURE LEGENDS

**Supplementary Figure 1 – Response and concordance of models.** A) Contingency table of patient response versus paired model. B) Distribution of patient response to treatment, based on paired model type. C) Model responsiveness to treatment between cancer types and D) treatment types. E) Comparison of concordance between patient and model across treatment types and F) all cancer types. ▲, N = 0; ●, concordance = 0; ▪, responsive = 0. P-values calculated with Fisher’s exact test. Abbreviations: PDX; patient-derived xenograft, PDO; patient-derived organoid, NSCLC; non-small cell lung cancer, SCLC; small cell lung cancer, CR; complete response, PD; progressive disease, PR; partial response, SD; stable disease.

**Supplementary Figure 2 – Predictive statistics comparing PDX and PDO for individual cancers with at least 3 patient-model pairs per model.** Sensitivity, Specificity, PPV and NPV between PDX and PDO for A) breast, B) pancreatic, C) NSCLC, D) colorectal, E) gastroesophageal and F) other GI cancers. Point estimates, 95% CI, and adjusted P values are shown. P-values were calculated with Fisher’s exact test and corrected to adjusted P-values using the Benjamini-Hochberg procedure. Abbreviations: PDX; patient-derived xenograft, PDO; patient-derived organoid, PPV; positive predictive value, NPV; negative predictive value, CI; confidence interval, NSCLC; non-small cell lung cancer, GI; gastrointestinal.

**Supplementary Figure 3 – Quality assessment heatmaps.** A) Risk of bias assessment of the individual patient-model pairs from included studies, and B) divided into PDX and PDO and sorted by reliability score. The 6-criterion score is adapted from the Newcastle-Ottawa scale:

1. Selection: Did the paper include treatment responses from all PDX- or PDO-matched patient pairs from the modeling project?
2. Ascertainment: Was the PDX or PDO successfully validated to originate from matching the expected human tissue of origin with a DNA, RNA, or protein-based assay, or by histology?
3. Patient Outcome: Was patient response determined by RECIST criteria, and was the PDX or PDO response data obtained objectively?
4. Patient Follow-Up: Was patient follow-up sufficiently long enough for meaningful survival times to be evaluated; a survival time of at least 6 months, or earlier in the event of earlier death?
5. Patient-Model Pair Treatment Reporting: Was the PDX or PDO-matched patient pair described with sufficient detail? This criterion is comprised of 4 sub-categories, each of which needed to be reported for the criterion to be scored:

1. Route of administration for the model is confirmed to be the same as for the patient.
2. Dosing regimen is described for the patient.
3. Dosing regimen is described for the PDX or PDO.
4. Previous lines of systemic treatment are described for the patient.
6. Reporting of Replicates: Were there at least n = 3 replicates for the PDX or PDO experiment?

Abbreviations: PDX; patient-derived xenograft, PDO; patient-derived organoid.

**Supplementary Figure 4 – Proportion of high reliable pairs per cancer.** Percent of high reliability patient-model pairs per cancer. ▲, N = 0 for cancer; ●, Reliability = 0 for cancer. P-values calculated with Fisher’s exact test. Abbreviations: HR; high reliability, PDX; patient-derived xenograft, PDO; patient-derived organoid, NSCLC; non-small cell lung cancer.

**Supplementary Figure 5 – Responsiveness in model is not associated with prolonged progression-free survival in matched patients for low reliability pairs.** Comparison of PFS in patients from low reliability patient-model pairs and from whom the associated model did or did not respond to treatment across A) entire cohort, B) PDX, and C) PDO. One sided P-values calculated using log-rank test (Methods). Below each Kaplan-Meier curve are multivariate Cox proportional hazard models assessing the association between variables and patient PFS after removing categories with insufficient sample size (Methods). Abbreviations: PDX; patient-derived xenograft, PDO; patient-derived organoid, NSCLC; non-small cell lung cancer, PFS; progression-free survival.

**Supplementary Figure 6 – Progression-free survival prediction based on PDX response, stratified by quality assessment metrics met.** Comparison of patient PFS separated on whether matched PDX model responded to treatment, given that patient-model pair met A-G) quality assessment metric 1 through 6. One sided P-values calculated using log-rank test (Methods). Below each Kaplan-Meier curve are multivariate Cox proportional hazard models assessing the association between variables and patient PFS after removing categories with insufficient sample size (Methods). Abbreviations: PDX; patient-derived xenograft, NSCLC; non-small cell lung cancer, PFS; progression-free survival, QA; quality assessment criteria met, N/A; not applicable-statistics not reported due to inadequate sample size.

**Supplementary Figure 7 – Progression-free survival prediction based on PDO response, stratified by quality assessment metrics met.** Comparison of patient PFS separated on whether matched PDO model responded to treatment, given that patient-model pair met A-G) quality assessment metric 1 through 6. There were no patient-PDO-model pairs that met criteria 5, sufficient reporting. One sided P-values calculated using log-rank test (Methods). Below each Kaplan-Meier curve are multivariate Cox proportional hazard models assessing the association between variables and patient PFS after removing categories with insufficient sample size (Methods). Abbreviations: PDO; patient-derived organoid, NSCLC; non-small cell lung cancer, PFS; progression-free survival, QA; quality assessment criteria met.

## SUPPLEMENTARY TABLE LEGENDS

**Supplementary Table 1 –** Response and concordance rates of PDX and PDO models across cancer-types. Abbreviations: PDX; patient-derived xenograft, PDO; patient-derived organoid, NSCLC; non-small cell lung cancer, SCLC; small cell lung cancer.

**Supplementary Table 2 – Cohort demographics across high versus low reliability patient-model pairs.** Demographic variables compared between high and low reliability patient-model pairs. For cancer type, the top six cancer types based on total N are shown. P-values were calculated using Fisher’s exact test. Adjusted P-values calculated using the Benjamini-Hochberg procedure. Abbreviations: PDX; patient-derived xenograft, PDO; patient-derived organoid, NSCLC; non-small cell lung cancer, NS; not significant.

**Supplementary Table 3 – Concordance segregated by clinical variables based on PDX versus PDO patient-model pairs for high reliability patient-model pairs**. Comparison of concordance across clinical variables, for the entire cohort, PDX, and PDO. For cancer type, the top six cancer types based on total N are shown. P-values were calculated using Fisher’s exact test. Adjusted P-values were calculated using the Benjamini-Hochberg procedure. Abbreviations: PDX; patient-derived xenograft, PDO; patient-derived organoid, NSCLC; non-small cell lung cancer, NS; not significant.

**Supplementary Table 4 – Concordance segregated by clinical variables based on PDX versus PDO patient-model pairs for low reliability patient-model pairs**. Comparison of concordance across clinical variables, for the entire cohort, PDX, and PDO. For cancer type, the top six cancer types based on total N are shown. P-values were calculated using Fisher’s exact test. Adjusted P-values were calculated using the Benjamini-Hochberg procedure. Abbreviations: PDX; patient-derived xenograft, PDO; patient-derived organoid, NSCLC; non-small cell lung cancer, NS; not significant.

## APPENDIX

1: Detailed search strategy.

2: Information on each included publication.

