## Supplemental Figure for "Comparative analysis of patient-derived organoids and patient-derived xenografts as avatar models for predicting response to anti-cancer therapy"

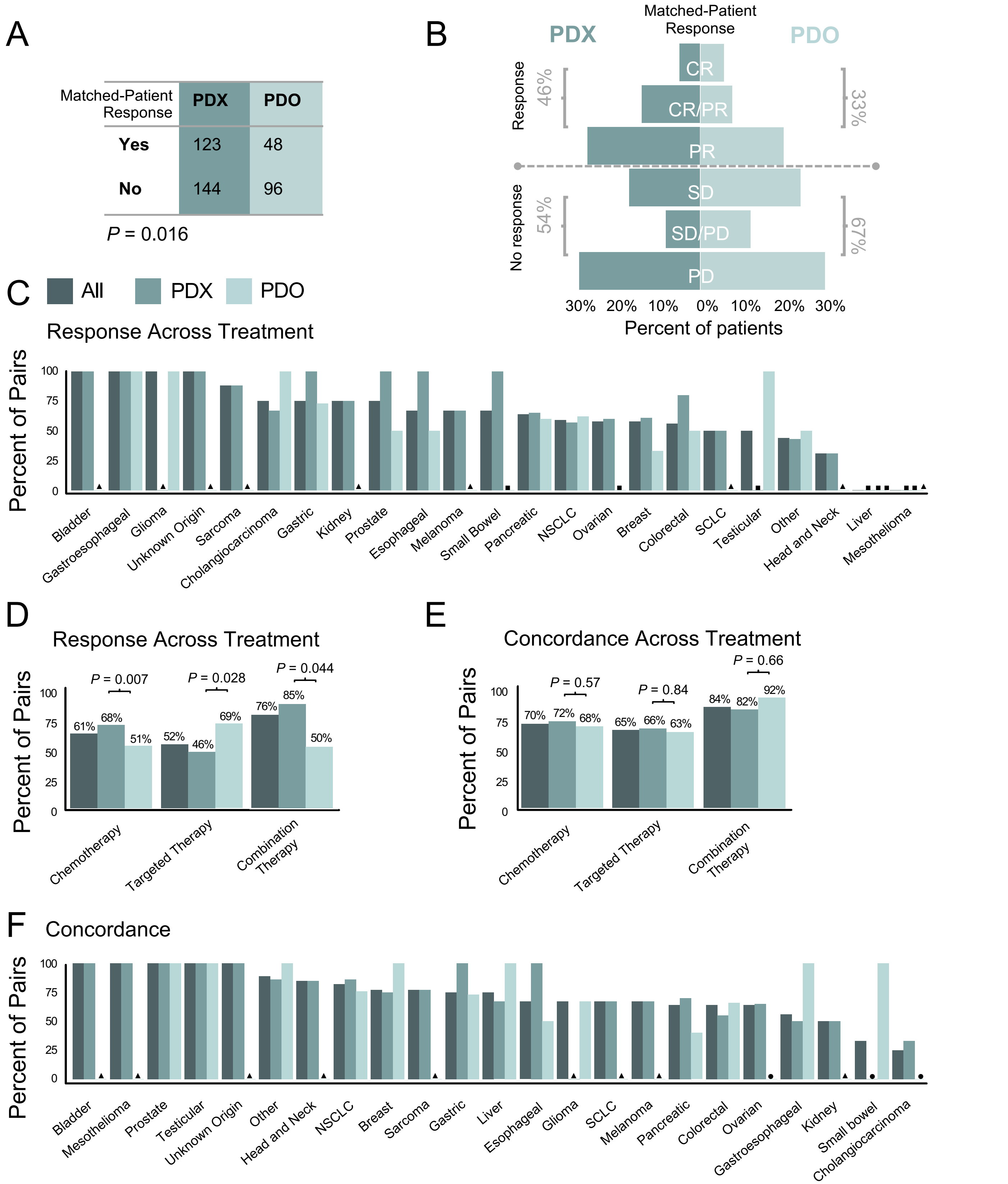


**Supplementary Figure 1 – Response and concordance of models.** A) Contingency table of patient response versus paired model. B) Distribution of patient response to treatment, based on paired model type. C) Model responsiveness to treatment between cancer types and D) treatment types. E) Comparison of concordance between patient and model across treatment types and F) all cancer types. ▲, N = 0; ●, concordance = 0; ■, responsive = 0. P-values calculated with Fisher's exact test. Abbreviations: PDX; patient-derived xenograft, PDO; patient-derived organoid, NSCLC; non-small cell lung cancer, SCLC; small cell lung cancer, CR; complete response, PD; progressive disease, PR; partial response, SD; stable disease.

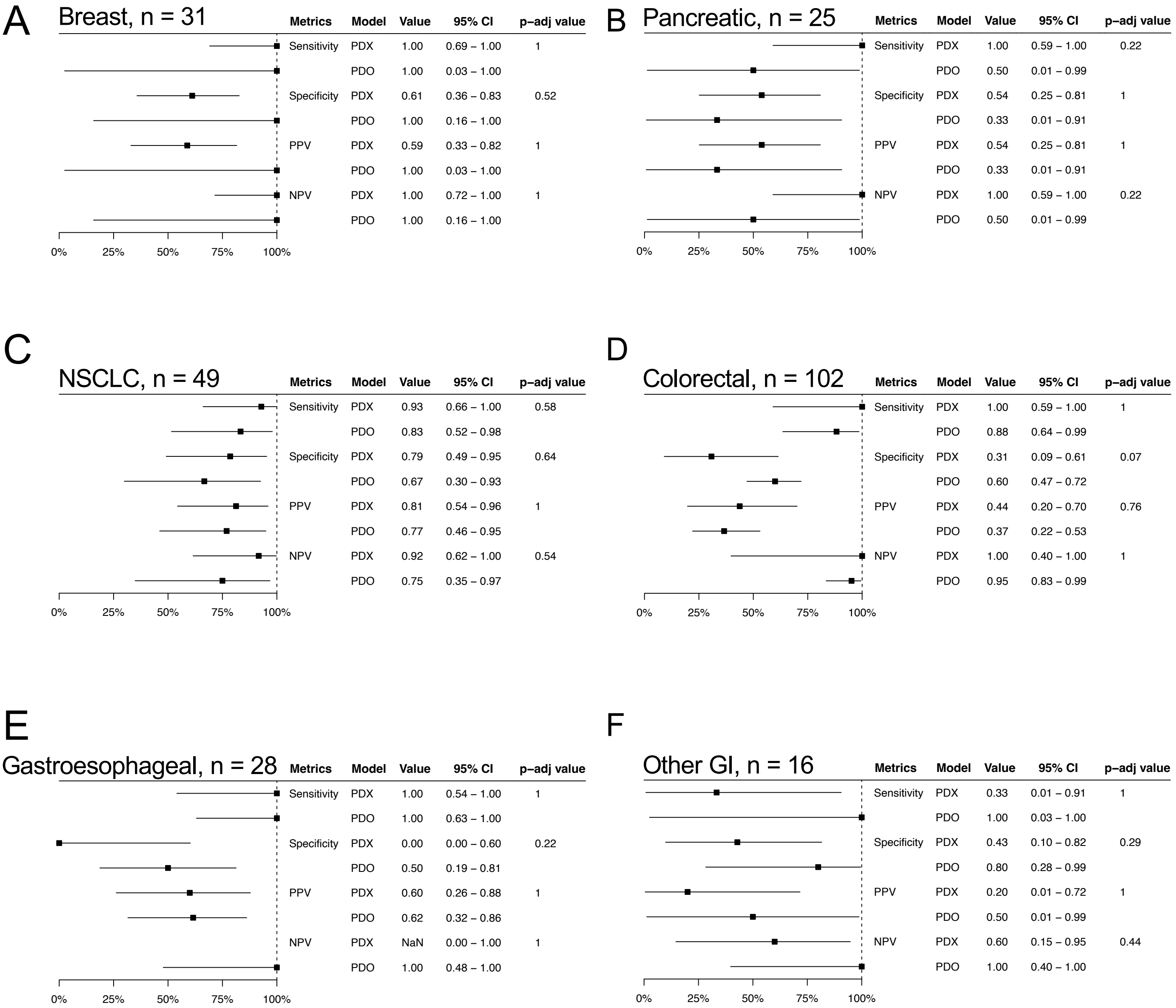


**Supplementary Figure 2 – Predictive statistics comparing PDX and PDO for individual cancers with at least 3 patient-model pairs per model.** Sensitivity, Specificity, PPV and NPV between PDX and PDO for A) breast, B) pancreatic, C) NSCLC, D) colorectal, E) gastroesophageal and F) other GI cancers. Point estimates, 95% CI, and adjusted P values are shown. P-values were calculated with Fisher's exact test and corrected to adjusted P-values using the Benjamini-Hochberg procedure. Abbreviations: PDX; patient-derived xenograft, PDO; patient-derived organoid, PPV; positive predictive value, NPV; negative predictive value, CI; confidence interval, NSCLC; non-small cell lung cancer, GI; gastrointestinal.

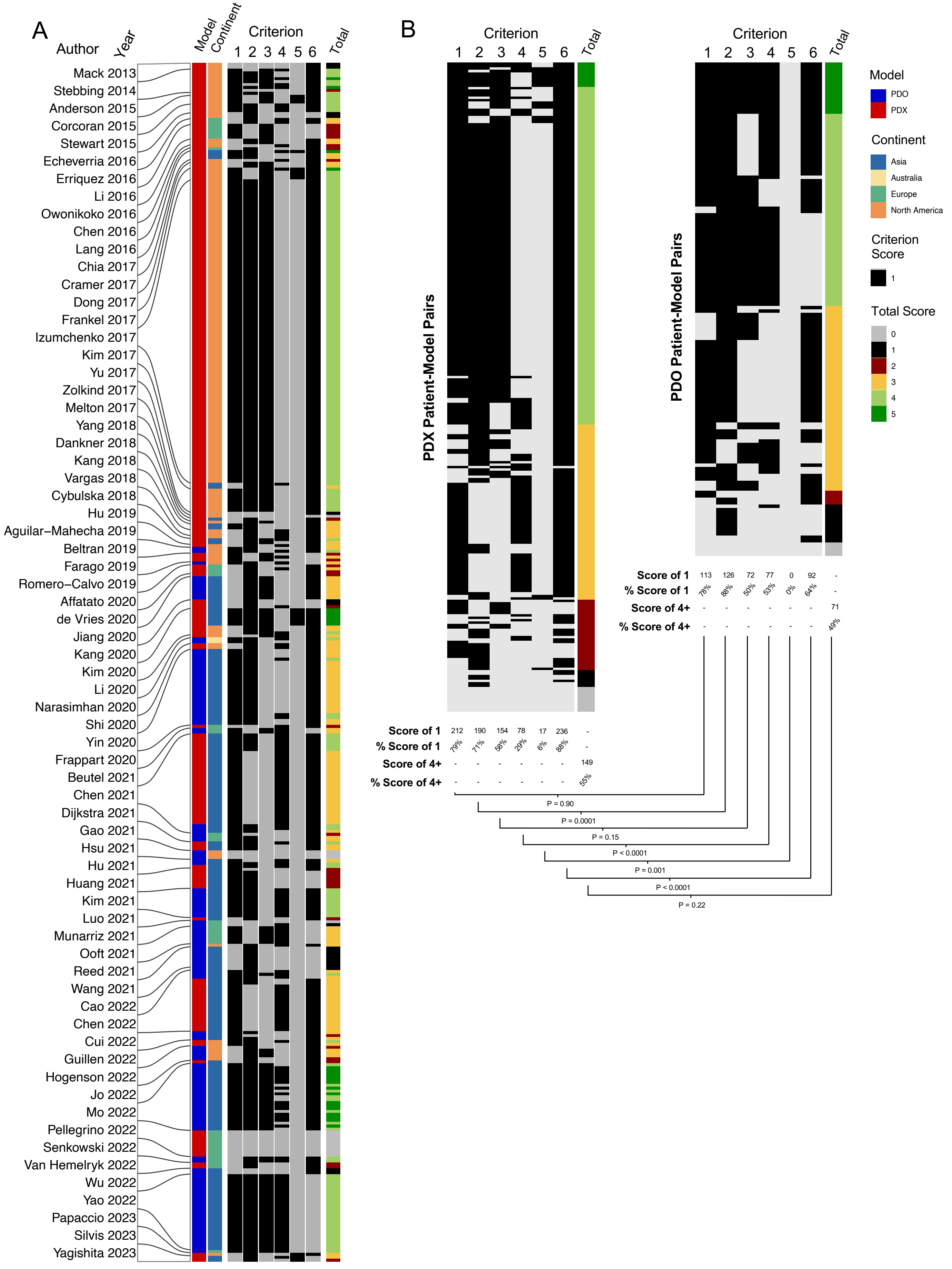
A
Author
Year
Model
Continent
Criterion
1 2 3 4 5 6
Total
Mack 2013
Stebbing 2014
Anderson 2015
Corcoran 2015
Stewart 2015
Echeverria 2016
Erriquez 2016
Li 2016
Owonikoko 2016
Chen 2016
Lang 2016
Chia 2017
Cramer 2017
Dong 2017
Frankel 2017
Izumchenko 2017
Kim 2017
Yu 2017
Zolkind 2017
Melton 2017
Yang 2018
Dankner 2018
Kang 2018
Vargas 2018
Cybulska 2018
Hu 2019
Aguilar–Mahecha 2019
Beltran 2019
Farago 2019
Romero–Calvo 2019
Affatato 2020
de Vries 2020
Jiang 2020
Kang 2020
Kim 2020
Li 2020
Narasimhan 2020
Shi 2020
Yin 2020
Frappart 2020
Beutel 2021
Chen 2021
Dijkstra 2021
Gao 2021
Hsu 2021
Hu 2021
Huang 2021
Kim 2021
Luo 2021
Munarriz 2021
Ooft 2021
Reed 2021
Wang 2021
Cao 2022
Chen 2022
Cui 2022
Guillen 2022
Hogenson 2022
Jo 2022
Mo 2022
Pellegrino 2022
Senkowski 2022
Van Hemelryk 2022
Wu 2022
Yao 2022
Papaccio 2023
Silvis 2023
Yagishita 2023
B
Criterion
1 2 3 4 5 6
Total
PDX Patient-Model Pairs
Score of 1: 212, 190, 154, 78, 17, 236, -
% Score of 1: 79%, 71%, 58%, 29%, 6%, 88%, -
Score of 4+: -, -, -, -, -, -, 149
% Score of 4+: -, -, -, -, -, -, 55%
PDO Patient-Model Pairs
Score of 1: 113, 126, 72, 77, 0, 92, -
% Score of 1: 78%, 88%, 50%, 53%, 0%, 64%, -
Score of 4+: -, -, -, -, -, -, 71
% Score of 4+: -, -, -, -, -, -, 49%
P = 0.90
P = 0.0001
P = 0.15
P < 0.0001
P = 0.001
P < 0.0001
P = 0.22
Model
PDO
PDX
Continent
Asia
Australia
Europe
North America
Criterion Score
1
Total Score
0
1
2
3
4
5

**Supplementary Figure 3 – Quality assessment heatmaps.** A) Risk-of-bias assessment of the individual patient-model pairs from included studies, and B) divided into PDX and PDO and sorted by reliability score. The 6-criterion score is adapted from the Newcastle-Ottawa scale:

1. Selection: Did the paper include treatment responses from all PDX- or PDO-matched patient pairs from the modeling project?
2. Ascertainment: Was the PDX or PDO successfully validated to originate from matching the expected human tissue of origin with a DNA, RNA, or protein-based assay, or by histology?
3. Patient Outcome: Was patient response determined by RECIST criteria, and was the PDX or PDO response data obtained objectively?
4. Patient Follow-Up: Was patient follow-up sufficiently long enough for meaningful survival times to be evaluated; a survival time of at least 6 months, or earlier in the event of earlier death?
5. Patient-Model Pair Treatment Reporting: Was the PDX or PDO-matched patient pair described with sufficient detail? This criterion is comprised of 4 sub-categories, each of which needed to be reported for the criterion to be scored:
    1. Route of administration for the model is confirmed to be the same as for the patient.
    2. Dosing regimen is described for the patient.
    3. Dosing regimen is described for the PDX or PDO.
    4. Previous lines of systemic treatment are described for the patient.
6. Reporting of Replicates: Were there at least n = 3 replicates for the PDX or PDO experiment?

Abbreviations: PDX; patient-derived xenograft, PDO; patient-derived organoid.

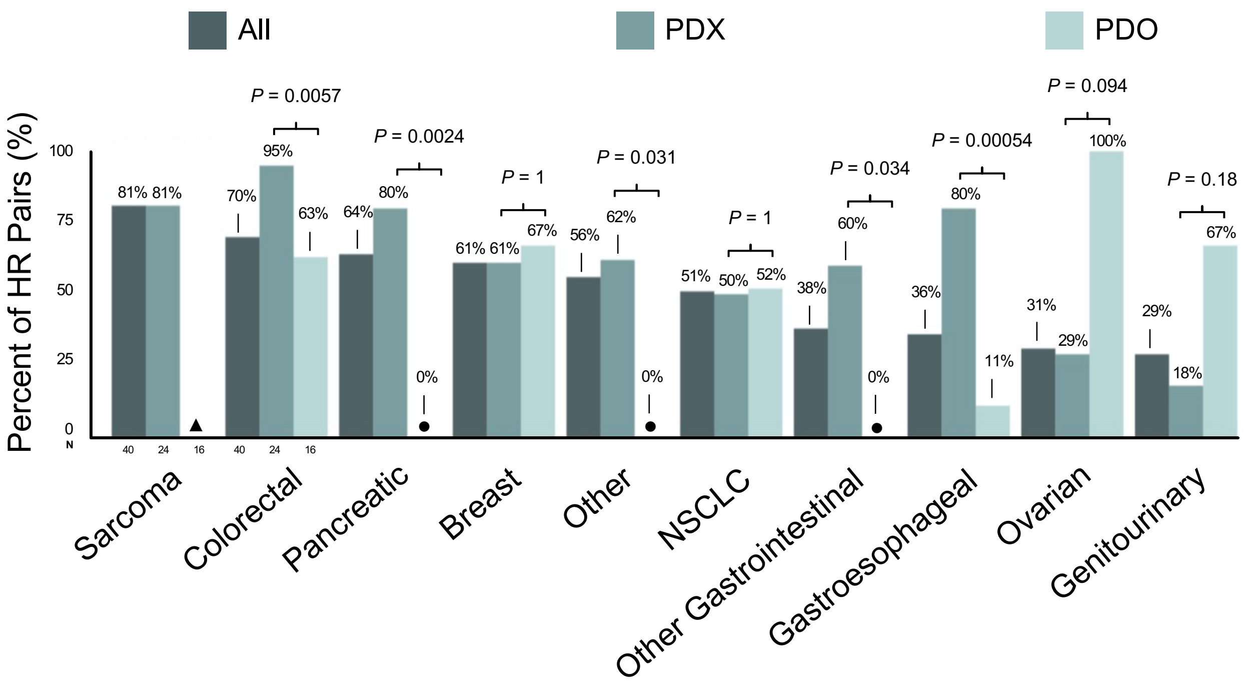


**Supplementary Figure 4 – Proportion of high reliable pairs per cancer.** Percent of high reliability patient-model pairs per cancer. ▲, N = 0 for cancer; ●, Reliability = 0 for cancer. P-values calculated with Fisher's exact test. Abbreviations: HR; high reliability, PDX; patient-derived xenograft, PDO; patient-derived organoid, NSCLC; non-small cell lung cancer.

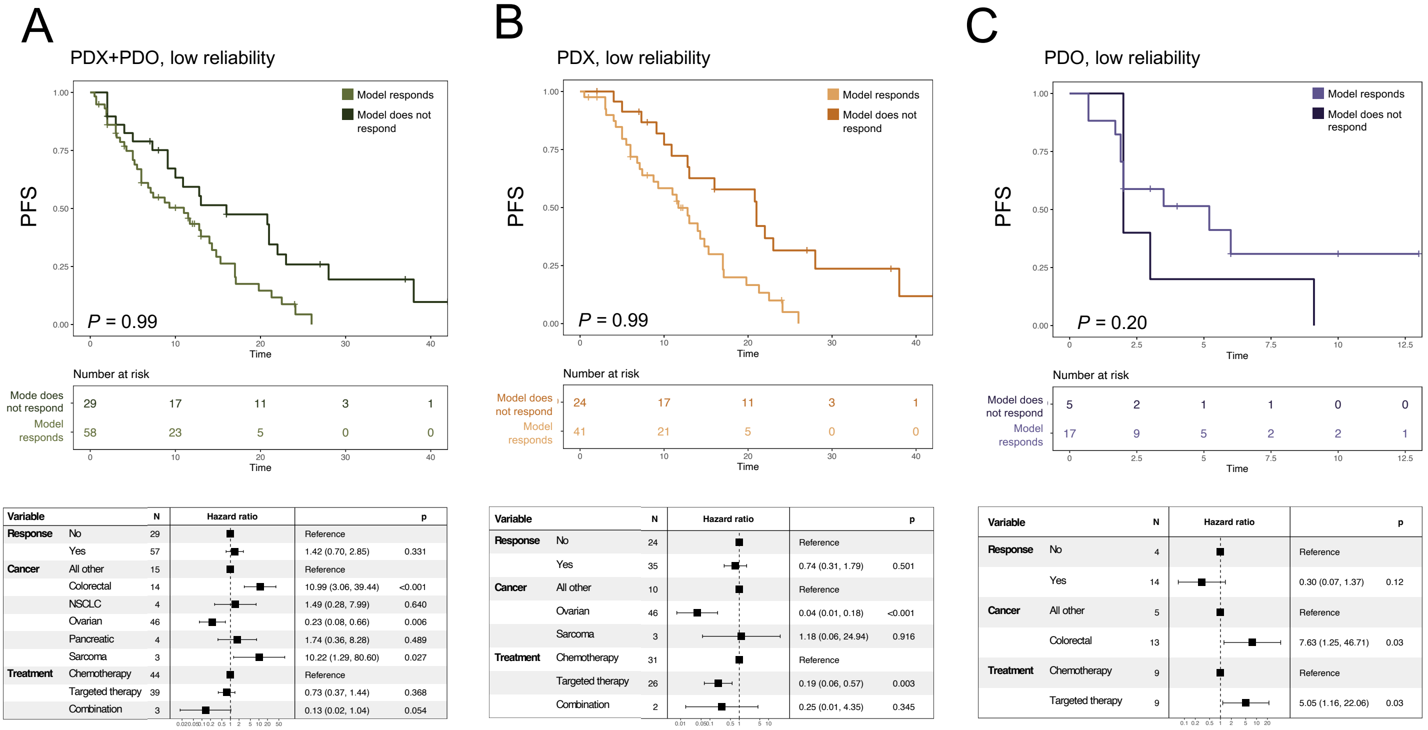


**Supplementary Figure 5 – Responsiveness in model is not associated with prolonged progression-free survival in matched patients for low reliability pairs.** Comparison of PFS in patients from low reliability patient-model pairs and from whom the associated model did or did not respond to treatment across A) entire cohort, B) PDX, and C) PDO. One sided P-values calculated using log-rank test (Methods). Below each Kaplan-Meier curve are multivariate Cox proportional hazard models assessing the association between variables and patient PFS after removing categories with insufficient sample size (Methods). Abbreviations: PDX; patient-derived xenograft, PDO; patient-derived organoid, NSCLC; non-small cell lung cancer, PFS; progression-free survival.

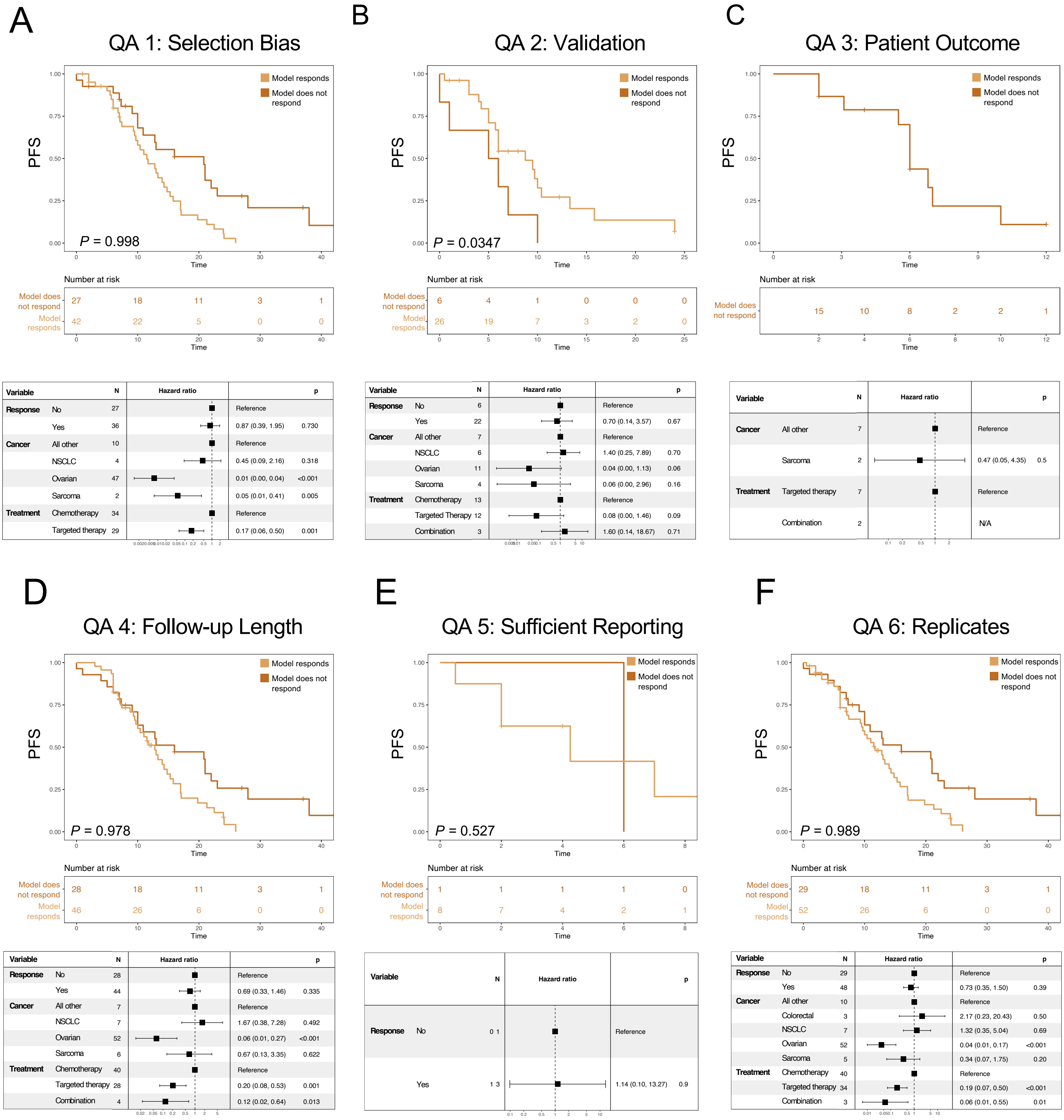


**Supplementary Figure 6 – Progression-free survival prediction based on PDX response, stratified by quality assessment metrics met.** Comparison of patient PFS separated on whether matched PDX model responded to treatment, given that patient-model pair met A-G) quality assessment metric 1 through 6. One sided P-values calculated using log-rank test (Methods). Below each Kaplan-Meier curve are multivariate Cox proportional hazard models assessing the association between variables and patient PFS after removing categories with insufficient sample size (Methods). Abbreviations: PDX; patient-derived xenograft, NSCLC; non-small cell lung cancer, PFS; progression-free survival, QA; quality assessment criteria met, N/A; not applicable-statistics not reported due to inadequate sample size.

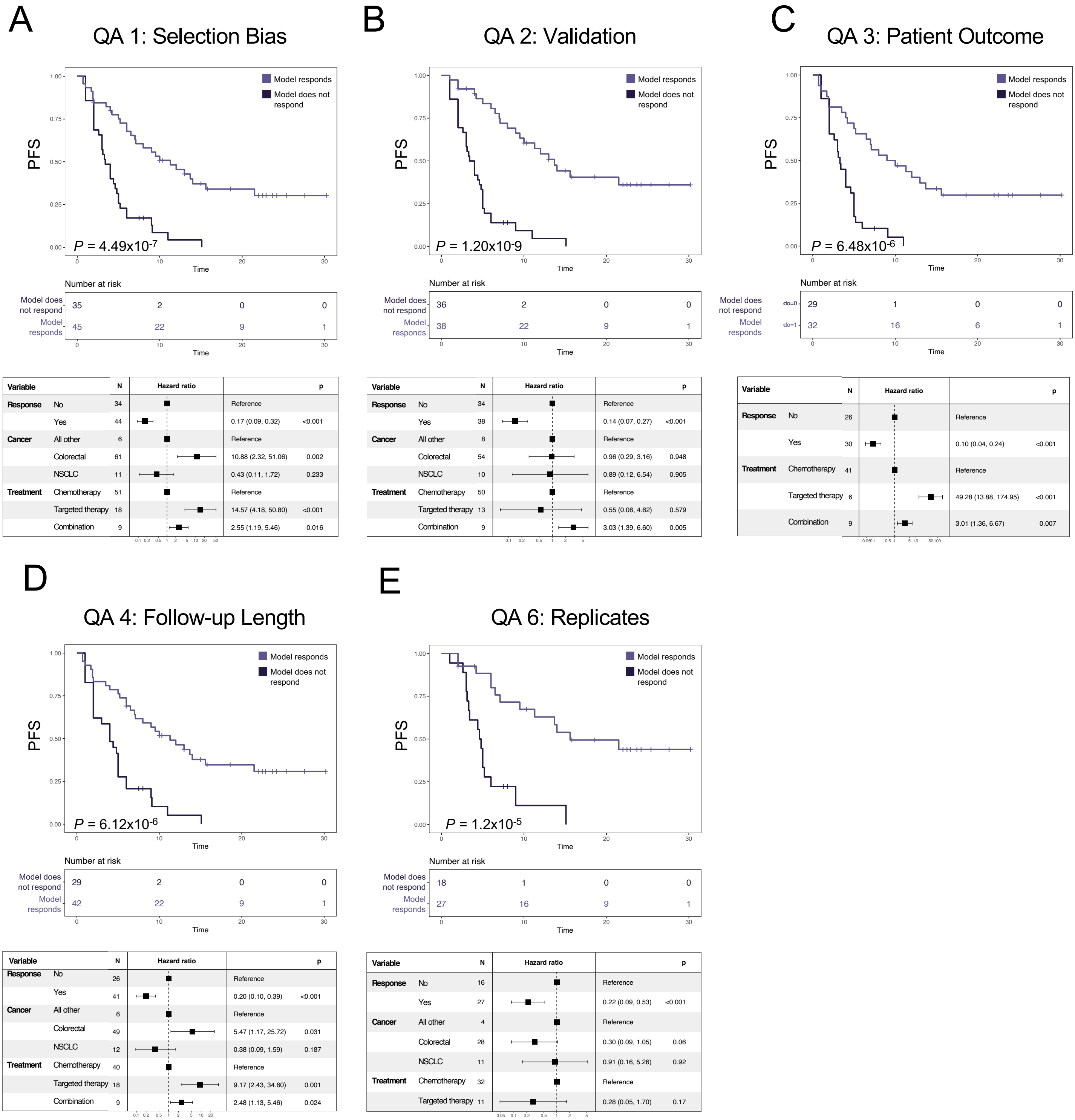


**Supplementary Figure 7 – Progression-free survival prediction based on PDO response, stratified by quality assessment metrics met.** Comparison of patient PFS separated on whether matched PDO model responded to treatment, given that patient-model pair met A-G) quality assessment metric 1 through 6. There were no patient-PDO-model pairs that met criteria 5, sufficient reporting. One sided P-values calculated using log-rank test (Methods). Below each Kaplan-Meier curve are multivariate Cox proportional hazard models assessing the association between variables and patient PFS after removing categories with insufficient sample size (Methods). Abbreviations: PDO; patient-derived organoid, NSCLC; non-small cell lung cancer, PFS; progression-free survival, QA; quality assessment criteria met.
