## Supplemental Tables for "Comparative analysis of patient-derived organoids and patient-derived xenografts as avatar models for predicting response to anti-cancer therapy"

| | Model Response | | | Concordance | | |
|---|---|---|---|---|---|---|
| | All | PDX | PDO | All | PDX | PDO |
| **Cancer** | | | | | | |
| Bladder cancer | 1/1 (100%) | 1/1 (100%) | - | 1/1 (100%) | 1/1 (100%) | - |
| Breast cancer | 18/31 (58%) | 17/28 (61%) | 1/3 (33%) | 24/31 (77%) | 21/28 (75%) | 3/3 (100%) |
| Cholangiocarcinoma | 3/4 (75%) | 2/3 (67%) | 1/1 (100%) | 1/4 (25%) | 1/3 (33%) | 0/1 (0%) |
| Colorectal cancer | 57/102 (56%) | 16/20 (80%) | 41/82 (50%) | 65/102 (64%) | 11/20 (55%) | 54/82 (66%) |
| Esophageal cancer | 2/3 (67%) | 1/1 (100%) | 1/2 (50%) | 2/3 (67%) | 1/1 (100%) | 1/2 (50%) |
| Gastric cancer | 12/16 (75%) | 1/1 (100%) | 11/15 (73%) | 12/16 (75%) | 1/1 (100%) | 11/15 (73%) |
| Gastroesophageal cancer | 9/9 (100%) | 8/8 (100%) | 1/1 (100%) | 5/9 (56%) | 4/8 (50%) | 1/1 (100%) |
| Glioma | 3/3 (100%) | - | 3/3 (100%) | 2/3 (67%) | - | 2/3 (67%) |
| Head and neck carcinoma | 4/13 (31%) | 4/13 (31%) | - | 11/13 (85%) | 11/13 (85%) | - |
| Kidney cancer | 3/4 (75%) | 3/4 (75%) | - | 2/4 (50%) | 2/4 (50%) | - |
| Liver cancer | 0/4 (0%) | 0/3 (0%) | 0/1 (0%) | 3/4 (75%) | 2/3 (67%) | 1/1 (100%) |
| NSCLC | 29/49 (59%) | 16/28 (57%) | 13/21 (62%) | 40/49 (82%) | 24/28 (86%) | 16/21 (76%) |
| SCLC | 3/6 (50%) | 3/6 (50%) | - | 4/6 (67%) | 4/6 (67%) | - |
| Melanoma | 4/6 (67%) | 4/6 (67%) | - | 4/6 (67%) | 4/6 (67%) | - |
| Mesothelioma | 0/4 (0%) | 0/4 (0%) | - | 4/4 (100%) | 4/4 (100%) | - |
| Other | 8/18 (44%) | 6/14 (43%) | 2/4 (50%) | 16/18 (89%) | 12/14 (86%) | 4/4 (100%) |
| Ovarian cancer | 45/77 (58%) | 45/75 (60%) | 0/2 (0%) | 49/77 (64%) | 49/75 (65%) | 0/2 (0%) |
| Pancreatic cancer | 16/25 (64%) | 13/20 (65%) | 3/5 (60%) | 16/25 (64%) | 14/20 (70%) | 2/5 (40%) |
| Prostate cancer | 3/4 (75%) | 2/2 (100%) | 1/2 (50%) | 4/4 (100%) | 2/2 (100%) | 2/2 (100%) |
| Sarcoma | 23/26 (88%) | 23/26 (88%) | - | 20/26 (77%) | 20/26 (77%) | - |
| Small bowel cancer | 2/3 (67%) | 2/2 (100%) | 0/1 (0%) | 1/3 (33%) | 0/2 (0%) | 1/1 (100%) |
| Testicular cancer | 1/2 (50%) | 0/1 (0%) | 1/1 (100%) | 2/2 (100%) | 1/1 (100%) | 1/1 (100%) |
| Unknown origin | 1/1 (100%) | 1/1 (100%) | - | 1/1 (100%) | 1/1 (100%) | - |

**Supplementary Table 1 – Response and concordance rates of PDX and PDO models across cancer-types.** Abbreviations: PDX; patient-derived xenograft, PDO; patient-derived organoid, NSCLC; non-small cell lung cancer, SCLC; small cell lung cancer.

| | High Reliability Cohort ( ≥ 4) | | | | Low Reliability Cohort ( < 4) | | | | High vs Low Reliability |
|---|---|---|---|---|---|---|---|---|---|
| | **Total** | **PDX** | **PDO** | **P-adj Value** | **Total** | **PDX** | **PDO** | **P-adj Value** | **P-adj Value** |
| **All** | 220 | 149 (68%) | 71 (32%) | - | 191 | 118 (62%) | 73 (38%) | - | - |
| **Continent** | | | | | | | | | |
| North America | 133 | 132 (99%) | 1 (1%) | < 0.00001 | 39 | 28 (72%) | 11 (28%) | NS | < 0.00001 |
| Asia | 83 | 17 (20%) | 66 (80%) | < 0.00001 | 114 | 66 (58%) | 48 (42%) | NS | < 0.00001 |
| Europe | 3 | 0 (0%) | 3 (100%) | NS | 37 | 24 (65%) | 13 (35%) | NS | NS |
| Australia | 1 | 0 (0%) | 1 (100%) | NS | 1 | 0 (0%) | 1 (100%) | NS | - |
| **Sex** | | | | | | | | | |
| Male | 101 | 60 (59%) | 29 (41%) | NS | 58 | 28 (48%) | 30 (52%) | NS | NS |
| Female | 89 | 60 (67%) | 40 (33%) | - | 41 | 27 (66%) | 14 (34%) | - | - |
| **Age, mean (min-max)** | | | | | | | | | |
| | 53.88 (6-83) | 51.99 (6-79) | 57.34 (24-83) | NS | 55.15 (1-77) | 53.66 (1-76) | 57.37 (18-77) | NS | NS |
| **Stage** | | | | | | | | | |
| Metastatic | 130 | 76 (58%) | 54 (42%) | NS | 102 | 60 (59%) | 42 (41%) | NS | NS |
| Non-metastatic | 31 | 24 (77%) | 4 (23%) | - | 32 | 22 (69%) | 10 (31%) | - | - |
| **Cancer** | | | | | | | | | |
| Colorectal | 71 | 19 (27%) | 52 (73%) | < 0.00001 | 31 | 1 (3%) | 30 (97%) | < 0.00001 | < 0.05 |
| Sarcoma | 21 | 21 (100%) | 0 (0%) | < 0.001 | 5 | 5 (100%) | 0 (0%) | NS | NS |
| NSCLC | 25 | 14 (56%) | 11 (44%) | NS | 24 | 14 (58%) | 10 (42%) | NS | NS |
| Ovarian | 24 | 22 (92%) | 2 (8%) | < 0.05 | 53 | 53 (100%) | 0 (0%) | < 0.00001 | NS |
| Breast | 19 | 17 (89%) | 2 (11%) | NS | 12 | 11 (92%) | 1 (8%) | NS | NS |
| Pancreatic | 16 | 16 (100%) | 0 (0%) | < 0.01 | 9 | 4 (44%) | 5 (56%) | NS | < 0.05 |
| Other | 44 | 40 (91%) | 4 (9%) | < 0.001 | 57 | 30 (53%) | 27 (47%) | NS | < 0.001 |
| **Treatment** | | | | | | | | | |
| Chemotherapy | 139 | 89 (64%) | 50 (36%) | NS | 105 | 58 (55%) | 47 (45%) | NS | NS |
| Targeted therapy | 46 | 34 (74%) | 12 (26%) | NS | 76 | 53 (70%) | 23 (30%) | NS | NS |
| Combination therapy | 35 | 26 (74%) | 9 (26%) | NS | 10 | 7 (70%) | 3 (30%) | NS | NS |

**Supplementary Table 2 – Cohort demographics across high versus low reliability patient-model pairs.** Demographic variables compared between high and low reliability patient-model pairs. For cancer type, the top six cancer types based on total N are shown. P-values were calculated using Fisher's exact test. Adjusted P-values calculated using the Benjamini-Hochberg procedure. Abbreviations: PDX; patient-derived xenograft, PDO; patient-derived organoid, NSCLC; non-small cell lung cancer, NS; not significant.

| | All – High Reliability Cohort | | | | PDX | | | | PDO | | | | PDX vs PDO |
|---|---|---|---|---|---|---|---|---|---|---|---|---|---|
| | Total | Concordant | Non-concordant | P-adj Value | Total | Concordant | Non-concordant | P-adj Value | Tot al | Concordan t | Non-concordant | P-adj Value | P-adj Value |
| **All** | 220 | 154 (70%) | 66 (30%) | - | 149 | 107 (72%) | 42 (28%) | - | 71 | 47 (66%) | 24 (34%) | NS | NS |
| **Continent** | | | | | | | | | | | | | |
| North America | 133 | 95 (71%) | 38 (29%) | NS | 132 | 94 (71%) | 38 (29%) | NS | 1 | 1 (100%) | 0 (0%) | NS | NS |
| Asia | 83 | 59 (71%) | 24 (29%) | NS | 17 | 13 (76%) | 4 (24%) | NS | 66 | 46 (70%) | 20 (30%) | NS | NS |
| Europe | 3 | 0 (0%) | 1 (100%) | NS | - | - | - | - | 3 | 0 (0%) | 3 (100%) | NS | - |
| Australia | 1 | 0 (0%) | 1 (100%) | NS | - | - | - | - | 1 | 0 (0%) | 1 (100%) | NS | - |
| **Sex** | | | | | | | | | | | | | |
| Male | 101 | 74 (73%) | 27 (27%) | NS | 60 | 45 (75%) | 15 (25%) | NS | 41 | 29 (71%) | 12 (29%) | NS | NS |
| Female | 89 | 56 (63%) | 33 (37%) | - | 60 | 39 (65%) | 21 (35%) | - | 29 | 17 (59%) | 12 (41%) | - | - |
| **Stage** | | | | | | | | | | | | | |
| Metastatic | 130 | 88 (68%) | 42 (32%) | NS | 76 | 52 (68%) | 24 (32%) | NS | 54 | 36 (67%) | 18 (33%) | NS | NS |
| Non-metastatic | 31 | 28 (90%) | 3 (10%) | - | 24 | 22 (92%) | 2 (8%) | - | 7 | 6 (86%) | 1 (14%) | - | - |
| **Cancer** | | | | | | | | | | | | | |
| Colorectal | 71 | 44 (62%) | 27 (38%) | NS | 19 | 11 (58%) | 8 (42%) | NS | 52 | 33 (63%) | 19 (37%) | NS | NS |
| Sarcoma | 21 | 15 (71%) | 6 (29%) | NS | 21 | 15 (71%) | 6 (29%) | NS | - | - | - | - | - |
| NSCLC | 25 | 21 (84%) | 4 (16%) | NS | 14 | 13 (93%) | 1 (7%) | NS | 11 | 8 (73%) | 3 (27%) | NS | NS |
| Ovarian | 24 | 16 (67%) | 8 (33%) | NS | 22 | 16 (73%) | 6 (27%) | NS | 2 | 0 (0%) | 1 (100%) | NS | NS |
| Breast | 19 | 16 (84%) | 3 (16%) | NS | 17 | 14 (82%) | 3 (18%) | NS | 2 | 2 (100%) | 0 (0%) | NS | NS |
| Pancreatic | 16 | 11 (69%) | 5 (31%) | NS | 16 | 11 (69%) | 5 (31%) | NS | - | - | - | - | - |
| Other | 44 | 31 (70%) | 13 (30%) | NS | 40 | 27 (68%) | 13 (32%) | NS | 4 | 4 (100%) | 0 (0%) | NS | NS |
| **Treatment** | | | | | | | | | | | | | |
| Chemotherapy | 139 | 93 (67%) | 46 (33%) | NS | 89 | 62 (70%) | 27 (30%) | NS | 50 | 31 (62%) | 19 (38%) | NS | NS |
| Targeted therapy | 46 | 31 (67%) | 15 (33%) | NS | 34 | 23 (68%) | 11 (32%) | NS | 12 | 8 (67%) | 4 (33%) | NS | NS |
| Combination therapy | 35 | 30 (86%) | 5 (14%) | NS | 26 | 22 (85%) | 4 (15%) | NS | 9 | 8 (89%) | 1 (11%) | NS | NS |

**Supplementary Table 3 – Concordance segregated by clinical variables based on PDX versus PDO patient-model pairs for high reliability patient-model pairs**. Comparison of concordance across clinical variables, for the entire cohort, PDX, and PDO. For cancer type, the top six cancer types based on total N are shown. P-values were calculated using Fisher's exact test. Adjusted P- values were calculated using the Benjamini-Hochberg procedure. Abbreviations: PDX; patient-derived xenograft, PDO; patient-derived organoid, NSCLC; non-small cell lung cancer, NS; not significant.

| | All – Low Reliability Cohort | | | | PDX | | | | PDO | | | | PDX vs PDO |
|---|---|---|---|---|---|---|---|---|---|---|---|---|---|
| | Total | Concordant | Non-concordant | P-adj Value | Total | Concordant | Non-concordant | P-adj Value | Total | Concordant | Non-concordant | P-adj Value | P-adj Value |
| **All** | 191 | 135 (71%) | 56 (29%) | - | 118 | 83 (70%) | 35 (30%) | - | 73 | 52 (71%) | 21 (29%) | NS | NS |
| **Continent** | | | | | | | | | | | | | |
| North America | 39 | 26 (67%) | 13 (22%) | NS | 28 | 20 (71%) | 8 (29%) | NS | 11 | 6 (55%) | 5 (45%) | NS | NS |
| Asia | 114 | 82 (72%) | 32 (28%) | NS | 66 | 42 (64%) | 24 (36%) | NS | 48 | 40 (83%) | 8 (17%) | NS | NS |
| Europe | 37 | 26 (70%) | 11 (30%) | NS | 24 | 21 (88%) | 3 (12%) | NS | 13 | 5 (38%) | 8 (62%) | NS | NS |
| Australia | 1 | 1 (100%) | 0 (0%) | NS | 0 | - | - | - | 1 | 1 (100%) | 0 (0%) | NS | - |
| **Sex** | | | | | | | | | | | | | |
| Male | 58 | 45 (78%) | 13 (22%) | NS | 28 | 23 (82%) | 5 (18%) | NS | 30 | 22 (73%) | 8 (27%) | NS | NS |
| Female | 41 | 32 (78%) | 9 (22%) | - | 27 | 19 (70%) | 8 (30%) | - | 14 | 13 (93%) | 1 (0.07%) | - | - |
| **Stage** | | | | | | | | | | | | | |
| Metastatic | 102 | 74 (73%) | 28 (27%) | NS | 60 | 44 (73%) | 16 (27%) | NS | 42 | 30 (71%) | 12 (29%) | NS | NS |
| Non-metastatic | 32 | 21 (66%) | 11 (34%) | - | 22 | 17 (77%) | 5 (23%) | - | 10 | 4 (40%) | 6 (60%) | - | - |
| **Cancer** | | | | | | | | | | | | | |
| Colorectal | 31 | 21 (68%) | 10 (32%) | NS | 1 | 0 (0%) | 1 (100%) | NS | 30 | 21 (70%) | 9 (30%) | NS | NS |
| Sarcoma | 5 | 5 (100%) | 0 (0%) | NS | 5 | 5 (100%) | 0 (0%) | NS | - | - | - | - | - |
| NSCLC | 24 | 19 (79%) | 5 (21%) | NS | 14 | 11 (79%) | 3 (21%) | NS | 10 | 8 (80%) | 2 (20%) | NS | NS |
| Ovarian | 53 | 33 (62%) | 20 (38%) | NS | 53 | 33 (62%) | 20 (38%) | NS | - | - | - | - | - |
| Breast | 12 | 8 (67%) | 4 (33%) | NS | 11 | 7 (64%) | 4 (36%) | NS | 1 | 1 (100%) | 0 (0%) | NS | NS |
| Pancreatic | 9 | 5 (56%) | 4 (44%) | NS | 4 | 3 (75%) | 1 (25%) | NS | 5 | 2 (40%) | 3 (60%) | NS | NS |
| Other | 57 | 44 (77%) | 13 (23%) | NS | 30 | 24 (80%) | 6 (20%) | NS | 27 | 20 (74%) | 7 (26%) | NS | NS |
| **Treatment** | | | | | | | | | | | | | |
| Chemotherapy | 105 | 79 (75%) | 26 (25%) | NS | 58 | 44 (76%) | 14 (24%) | NS | 47 | 35 (74%) | 12 (26%) | NS | NS |
| Targeted therapy | 76 | 48 (63%) | 28 (37%) | NS | 53 | 34 (64%) | 19 (36%) | NS | 23 | 14 (61%) | 9 (39%) | NS | NS |
| Combination therapy | 10 | 8 (80%) | 2 (20%) | NS | 7 | 5 (71%) | 2 (29%) | NS | 3 | 3 (100%) | 0 (0%) | NS | NS |

**Supplementary Table 4 – Concordance segregated by clinical variables based on PDX versus PDO patient-model pairs for low reliability patient-model pairs**. Comparison of concordance across clinical variables, for the entire cohort, PDX, and PDO. For cancer type, the top six cancer types based on total N are shown. P-values were calculated using Fisher's exact test. Adjusted P- values were calculated using the Benjamini-Hochberg procedure. Abbreviations: PDX; patient-derived xenograft, PDO; patient-derived organoid, NSCLC; non-small cell lung cancer, NS; not significant.
