## Appendices for "Comparative analysis of patient-derived organoids and patient-derived xenografts as avatar models for predicting response to anti-cancer therapy"

MEDLINE (via PubMed):

"pdx" OR
"patient-derived xenograft*" OR
"patient derived xenograft*" OR
"pdo" OR
"patient-derived organoid*" OR
"patient derived organoid*" OR
"organoid*" AND
("cancer" OR "melanoma" OR "glio*" OR "tumor*" OR "tumour*")

EMBASE (via OVID):

(("pdx" or "patient-derived xenograft*" or "patient derived xenograft*" or "pdo" or "patient-derived organoid*" or "patient derived organoid*" or "organoid*") and ("cancer" or "melanoma" or "glio*" or "tumor*" or "tumour*")).mp. [mp=title, book title, abstract, original title, name of substance word, subject heading word, floating sub-heading word, keyword heading word, organism supplementary concept word, protocol supplementary concept word, rare disease supplementary concept word, unique identifier, synonyms]

**Appendix 1 – Detailed search strategy.**

| Paper | Journal | DOI or Permalink | Year | Lead Author | Paper or Abstract | Method of Acquisition | Model Type | Number of Patient-Model Pairs | Primary Tumor Type(s) |
|---|---|---|---|---|---|---|---|---|---|
| Divergent activity of afatinib (AFAT) and cetuximab (CET) in patient-derived xenograft (PDX) models of acquired erlotinib resistance | Journal of Thoracic Oncology | https://escholarship.org/uc/item/86k7m2wr | 2013 | Mack | Abstract | Search Strategy | PDX | 2 | NSCLC |
| Patient-derived xenografts for individualized care in advanced sarcoma | Cancer | 10.1002/cncr.28696 | 2014 | Stebbing | Paper | Search Strategy | PDX | 8 | Sarcoma |
| Initiation and characterization of small cell lung cancer patient-derived xenografts from ultrasound-guided transbronchial needle aspirates | PLOS ONE | 10.1371/journal.pone.0125255 | 2015 | Anderson | Paper | Search Strategy | PDX | 1 | Other - Lung Cancer (SCLC) |
| Combined BRAF and MEK Inhibition With Dabrafenib and Trametinib in BRAF V600–Mutant Colorectal Cancer | Journal of Clinical Oncology | 10.1200/JCO.2015.63.2471 | 2015 | Corcoran | Paper | Search Strategy | PDX | 3 | Colorectal Cancer |
| Clinical Utility of Patient-Derived Xenografts to Determine Biomarkers of Prognosis and Map Resistance Pathways in EGFR-Mutant Lung Adenocarcinoma | Journal of Clinical Oncology | 10.1200/JCO.2014.60.1492 | 2015 | Stewart | Paper | Search Strategy | PDX | 3 | NSCLC |
| Identifying and targeting chemoresistant subclones in triple negative breast cancer | Cancer Research | 10.1158/1538-7445.AM2016-2406 | 2016 | Echeverria | Abstract | Search Strategy | PDX | 2 | Breast Cancer |
| Xenopatients show the need for precision medicine approach to chemotherapy in ovarian cancer | Oncotarget | 10.18632/oncotarget.8325 | 2016 | Erriquez | Abstract | Search Strategy | PDX | 2 | Ovarian Cancer |
| Comparing Patient-Derived Xenograft and Computational Response Prediction for Targeted Therapy in Patients of Early-Stage Large Cell Lung Cancer | Clinical Cancer Research | 10.1158/1078-0432.CCR-15-2401 | 2016 | Li | Paper | Search Strategy | PDX | 5 | NSCLC |
| Patient-derived xenografts faithfully replicated clinical outcome in a phase II co-clinical trial of arsenic trioxide in relapsed small cell lung cancer | Journal of Translational Medicine | 10.1186/s12967-016-0861-5 | 2016 | Owonikoko | Paper | Search Strategy | PDX | 2 | Other - Lung Cancer (SCLC) |
| Targeting renal cell carcinoma with a HIF-2 antagonist | Nature | 10.1038/nature19796 | 2016 | Chen | Paper | Author Acquired | PDX | 1 | Genitourinary Cancer |
| Establishment of a large panel of patient-derived preclinical models of human renal cell carcinoma | Oncotarget | 10.18632/oncotarget.10659 | 2016 | Lang | Paper | Author Acquired | PDX | 1 | Genitourinary Cancer |
| Phenotype-driven precision oncology as a guide for clinical decisions one patient at a time | Nature Communications | 10.1038/s41467-017-00451-5 | 2017 | Chia | Paper | Search Strategy | PDX | 3 | Gastroesophageal Cancer, Other - Head and Neck Carcinoma |
| Pediatric Anaplastic Embryonal Rhabdomyosarcoma: Targeted Therapy Guided by Genetic Analysis and a Patient-Derived Xenograft Study | Frontiers in Oncology | 10.3389/fonc.2017.00327 | 2017 | Cramer | Paper | Search Strategy | PDX | 1 | Sarcoma |
| Tumor Xenografts of Human Clear Cell Renal Cell Carcinoma But Not Corresponding Cell Lines Recapitulate Clinical Response to Sunitinib: Feasibility of Using Biopsy Samples | European Urology Focus | 10.1016/j.euf.2016.08.005 | 2017 | Dong | Paper | Search Strategy | PDX | 2 | Genitourinary Cancer |
| Digoxin Plus Trametinib Therapy Achieves Disease Control in BRAF Wild-Type Metastatic Melanoma Patients | Neoplasia | 10.1016/j.neo.2017.01.010 | 2017 | Frankel | Paper | Search Strategy | PDX | 4 | Other - Melanoma |
| Patient-derived xenografts effectively capture responses to oncology therapy in a heterogeneous cohort of patients with solid tumors | Annals of Oncology | 10.1093/annonc/mdx416 | 2017 | Izumchenko | Paper | Search Strategy | PDX | 104 | Sarcoma, NSCLC, Colorectal Cancer, Gastroesophageal Cancer, Other Gastrointestinal Cancer, Breast Cancer, Pancreatic Cancer, Ovarian Cancer, Genitourinary Cancer, Other |
| Co-clinical trials demonstrate predictive biomarkers for dovitinib, an FGFR inhibitor, in lung squamous cell carcinoma | Annals of Oncology | 10.1093/annonc/mdx098 | 2017 | Kim | Paper | Search Strategy | PDX | 2 | NSCLC |
| Establishing and characterizing patient-derived xenografts using pre-chemotherapy percutaneous biopsy and post-chemotherapy surgical samples from a prospective neoadjuvant breast cancer study | Breast Cancer Research | 10.1186/s13058-017-0920-8 | 2017 | Yu | Paper | Search Strategy | PDX | 8 | Breast Cancer |
| Genomic and functional correlates from a phase II clinical trial of trametinib in surgically resectable oral cavity squamous cell carcinoma | Clinical Cancer Research | 10.1158/1557-3265.AACRAHNS17-PR01 | 2017 | Zolkind | Abstract | Search Strategy | PDX | 1 | Other - Head and Neck Carcinoma |
| A new ER+ Her2- Palbociclib resistant breast cancer PDX model | Cancer Research | 10.1158/1538-7445.AM2021-2994 | 2017 | Melton | Abstract | Search Strategy | PDX | 1 | Breast Cancer |
| Afatinib treatment for Her-2 amplified metastatic colorectal cancer based on patient-derived xenograft models and next generation sequencing | Cancer Biology & Therapy | 10.1080/15384047.2018.1529120 | 2018 | Yang | Paper | Search Strategy | PDX | 1 | Colorectal Cancer |
| Dual MAPK Inhibition Is an Effective Therapeutic Strategy for a Subset of Class II BRAF Mutant Melanomas | Clinical Cancer Research | 10.1158/1078-0432.CCR-17-3384 | 2018 | Dankner | Paper | Search Strategy | PDX | 1 | Other - Melanoma |
| Establishment of a platform of non-small-cell lung cancer patient-derived xenografts with clinical and genomic annotation | Lung Cancer | 10.1016/j.lungcan.2018.08.008 | 2018 | Kang | Paper | Search Strategy | PDX | 2 | NSCLC |
| Case study: patient-derived clear cell adenocarcinoma xenograft model longitudinally predicts treatment response | npj Precision Oncology | 10.1038/s41698-018-0060-3 | 2018 | Vargas | Paper | Search Strategy | PDX | 2 | Ovarian Cancer |
| A Genomically Characterized Collection of High-Grade Serous Ovarian Cancer Xenografts for Preclinical Testing | American Journal of Pathology | 10.1016/j.ajpath.2018.01.019 | 2018 | Cybulska | Paper | Author Acquired | PDX | 1 | Ovarian Cancer |
| Establishment of a hepatocellular carcinoma patient-derived xenograft platform and its application in biomarker identification | International Journal of Cancer | 10.1002/ijc.32564 | 2019 | Hu | Paper | Search Strategy | PDX | 2 | Other Gastrointestinal Cancer |
| Precision Medicine Tools to Guide Therapy and Monitor Response to Treatment in a HER-2+ Gastric Cancer Patient: Case Report | Frontiers in Oncology | 10.3389/fonc.2019.00698 | 2019 | Aguilar-Mahecha | Paper | Search Strategy | PDX | 1 | Gastroesophageal Cancer |
| A Phase II Trial of the Aurora Kinase A Inhibitor Alisertib for Patients with Castration-resistant and Neuroendocrine Prostate Cancer: Efficacy and Biomarkers | Clinical Cancer Research | 10.1158/1078-0432.CCR-18-1912 | 2019 | Beltran | Paper | Search Strategy | PDO | 2 | Genitourinary Cancer |
| Combination olaparib and temozolomide in relapsed small-cell lung cancer | Cancer Discovery | 10.1158/2159-8290.CD-19-0582 | 2019 | Farago | Paper | Search Strategy | PDX | 3 | Other - Lung Cancer (SCLC) |
| Human organoids share structural and genetic features with primary pancreatic adenocarcinoma tumors | Molecular Cancer Research | 10.1158/1541-7786.MCR-18-0531 | 2019 | Romero-Calvo | Paper | Search Strategy | PDO | 1 | Pancreatic Cancer |
| Establishment and Characterization of Patient-Derived Xenografts (PDXs) of Different Histology from Malignant Pleural Mesothelioma Patients | Cancers | 10.3390/cancers12123846 | 2020 | Affatato | Paper | Search Strategy | PDX | 3 | Other - Mesothelioma |
| Establishment and characterisation of testicular cancer patient-derived xenograft models for preclinical evaluation of novel therapeutic strategies | Scientific Reports | 10.1038/s41598-020-75518-3 | 2020 | de Vries | Paper | Search Strategy | PDX | 1 | Genitourinary Cancer |
| An Automated Organoid Platform with Inter-organoid Homogeneity and Inter-patient Heterogeneity | Cell Reports Medicine | 10.1016/j.xcrm.2020.100161 | 2020 | Jiang | Paper | Search Strategy | PDO | 8 | Colorectal Cancer, Gastroesophageal Cancer, Other Gastrointestinal Cancer |
| Establishment and characterization of patient-derived xenografts as paraclinical models for head and neck cancer | BMC Cancer | 10.1186/s12885-020-06786-5 | 2020 | Kang | Paper | Search Strategy | PDX | 3 | Other - Head and Neck Carcinoma |
| Mouse-human co-clinical trials demonstrate superior anti-tumour effects of buparlisib (BKM120) and cetuximab combination in squamous cell carcinoma of head and neck | British Journal of Cancer | 10.1038/s41416-020-01074-2 | 2020 | Kim | Paper | Search Strategy | PDX | 6 | Other - Head and Neck Carcinoma |
| Modeling biological and genetic diversity in upper tract urothelial carcinoma with patient derived xenografts | Nature Communications | 10.1038/s41467-020-15885-7 | 2020 | Kim | Paper | Search Strategy | PDX | 3 | Genitourinary Cancer |

**Appendix 2 – Information on each included publication.**

| Paper | Journal | DOI or Permalink | Year | Lead Author | Paper or Abstract | Method of Acquisition | Model Type | Number of Patient-Model Pairs | Primary Tumor Type(s) |
|---|---|---|---|---|---|---|---|---|---|
| Her2-mediated internalization of cytotoxic agents in ERBB2 amplified or mutant lung cancers | Cancer Discovery | 10.1158/2159-8290.CD-20-0215 | 2020 | Li | Paper | Search Strategy | PDX | 1 | NSCLC |
| Medium-throughput Drug Screening of Patient-derived Organoids from Colorectal Peritoneal Metastases to Direct Personalized Therapy | Clinical Cancer Research | 10.1158/1078-0432.CCR-20-0073 | 2020 | Narasimhan | Paper | Search Strategy | PDO | 2 | Colorectal Cancer |
| BRAF V600E mutation and MET amplification as resistance pathways of the second-generation anaplastic lymphoma kinase (ALK) inhibitor alectinib in lung cancer | Lung Cancer | 10.1016/j.lungcan.2020.05.018 | 2020 | Shi | Paper | Search Strategy | PDX | 2 | NSCLC |
| Patient-derived tumor-like cell clusters for drug testing in cancer therapy | Science Translational Medicine | 10.1126/scitranslmed.aaz1723 | 2020 | Yin | Paper | Search Strategy | PDO | 26 | Colorectal Cancer, Gastroesophageal Cancer |
| Pancreatic cancer-derived organoids - a disease modeling tool to predict drug response | United European Gastroenterology Journal | 10.1177/2050640620905183 | 2020 | Frappart | Paper | Author Acquired | PDX | 1 | Pancreatic Cancer |
| A Prospective Feasibility Trial to Challenge Patient–Derived Pancreatic Cancer Organoids in Predicting Treatment Response | Cancers | 10.3390/cancers13112539 | 2021 | Beutel | Paper | Search Strategy | PDO | 2 | Pancreatic Cancer |
| Patient-Derived Xenografts Are a Reliable Preclinical Model for the Personalized Treatment of Epithelial Ovarian Cancer | Frontiers in Oncology | 10.3389/fonc.2021.744256 | 2021 | Chen | Paper | Search Strategy | PDX | 31 | Ovarian Cancer |
| Patient-Derived Organoids Can Guide Personalized-Therapies for Patients with Advanced Breast Cancer | Advanced Science | 10.1002/advs.202101176 | 2021 | Chen | Paper | Search Strategy | PDO | 3 | Breast Cancer |
| Patient-Derived Organoid Models of Human Neuroendocrine Carcinoma | Frontiers in Endocrinology | 10.3389/fendo.2021.627819 | 2021 | Dijkstra | Paper | Search Strategy | PDO | 3 | Other Gastrointestinal Cancer |
| Patient-Derived Xenograft Models for Intrahepatic Cholangiocarcinoma and Their Application in Guiding Personalized Medicine | Frontiers in Oncology | 10.3389/fonc.2021.704042 | 2021 | Gao | Paper | Search Strategy | PDX | 3 | Other Gastrointestinal Cancer |
| Micro-organospheres as a novel precision oncology platform in colorectal cancer | Annals of Oncology | 10.1016/j.annonc.2021.08.1725 | 2021 | Hsu | Abstract | Search Strategy | PDO | 3 | Colorectal Cancer |
| Lung cancer organoids analyzed on microwell arrays predict drug responses of patients within a week | Nature Communications | 10.1038/s41467-021-22676-1 | 2021 | Hu | Paper | Search Strategy | PDX & PDO | 3 | NSCLC |
| Predicting HCC Response to Multikinase Inhibitors With In Vivo Cirrhotic Mouse Model for Personalized Therapy | Cellular and Molecular Gastroenterology and Hepatology | 10.1016/j.jcmgh.2020.12.009 | 2021 | Huang | Paper | Search Strategy | PDX | 1 | Other Gastrointestinal Cancer |
| A Novel, Personalized Drug-Screening System for Platinum-Resistant Ovarian Cancer Patients: A Preliminary Clinical Report | Cancer Management and Research | 10.2147/CMAR.S276799 | 2021 | Huang | Paper | Search Strategy | PDX | 6 | Ovarian Cancer |
| Modeling Clinical Responses to Targeted Therapies by Patient-Derived Organoids of Advanced Lung Adenocarcinoma | Clinical Cancer Research | 10.1158/1078-0432.CCR-20-5026 | 2021 | Kim | Paper | Search Strategy | PDO | 10 | NSCLC |
| Establishment and characterization of an ovarian yolk sac tumor patient-derived xenograft model | Pediatric Surgery International | 10.1007/s00383-021-04895-1 | 2021 | Luo | Paper | Search Strategy | PDX | 1 | Ovarian Cancer |
| Patient-derived lung cancer organoids for the selection of therapeutic options in an ALK-rearranged tumor | Journal of Clinical Oncology | 10.1200/JCO.2021.39.15_suppl.e21014 | 2021 | Munarriz | Abstract | Search Strategy | PDO | 2 | NSCLC |
| Prospective experimental treatment of colorectal cancer patients based on organoid drug responses | ESMO Open | 10.1016/j.esmoop.2021.100103 | 2021 | Ooft | Paper | Search Strategy | PDO | 6 | Colorectal Cancer |
| A Functional Precision Medicine Pipeline Combines Comparative Transcriptomics and Tumor Organoid Modeling to Identify Bespoke Treatment Strategies for Glioblastoma | Cells | 10.3390/cells10123400 | 2021 | Reed | Paper | Search Strategy | PDO | 1 | Other - Glioma |
| Malignant effusion-derived non-small cell lung cancer organoid might be a feasible in vitro model for therapeutic screening | Annals of Oncology | 10.1016/j.annonc.2021.08.1985 | 2021 | Wang | Abstract | Search Strategy | PDO | 7 | NSCLC |
| Patient-Derived Organoid Facilitating Personalized Medicine in Gastrointestinal Stromal Tumor With Liver Metastasis: A Case Report | Frontiers in Oncology | 10.3389/fonc.2022.920762 | 2022 | Cao | Paper | Search Strategy | PDO | 1 | Other - Gastrointestinal Stromal Tumor |
| Patient-derived tumor organoids as a platform of precision treatment for malignant brain tumors | Scientific Reports | 10.1038/s41598-022-20487-y | 2022 | Chen | Paper | Search Strategy | PDO | 3 | Genitourinary Cancer, Other - Glioma |
| Generation and characterization of patient-derived xenografts from patients with osteosarcoma | Tissue and Cell | 10.1016/j.tice.2022.101911 | 2022 | Chen | Paper | Search Strategy | PDX | 2 | Sarcoma |
| Using Patient-Derived Xenograft (PDX) Models as a 'Black Box' to Identify More Applicable Patients for ADP-Ribose Polymerase Inhibitor (PARPi) Treatment in Ovarian Cancer: Searching for Novel Molecular and Clinical Biomarkers and Performing a Prospective Preclinical Trial | Cancers | 10.3390/cancers14194649 | 2022 | Chen | Paper | Search Strategy | PDX | 16 | Ovarian Cancer |
| Establishment of organoid models based on a nested array chip for fast and reproducible drug testing in colorectal cancer therapy | Bio-Design and Manufacturing | 10.3390/cancers14194649 | 2022 | Cui | Abstract | Search Strategy | PDO | 3 | Colorectal Cancer |
| A human breast cancer-derived xenograft and organoid platform for drug discovery and precision oncology | Nature Cancer | 10.1038/s43018-022-00337-6 | 2022 | Guillen | Paper | Search Strategy | PDX | 2 | Breast Cancer |
| Culture media composition influences patient-derived organoids ability to predict therapeutic response in gastrointestinal cancers | JCI Insight | 10.1172/jci.insight.158060 | 2022 | Hogenson | Paper | Search Strategy | PDO | 5 | Colorectal Cancer, Other Gastrointestinal Cancer |
| Comparative Study on the Efficacy and Exposure of Molecular Target Agents in Non-small Cell Lung Cancer PDX Models with Driver Genetic Alterations | Molecular Cancer Therapeutics | 10.1158/1535-7163.MCT-21-0371 | 2022 | Jo | Paper | Search Strategy | PDX | 1 | NSCLC |
| Patient-Derived Organoids from Colorectal Cancer with Paired Liver Metastasis Reveal Tumor Heterogeneity and Predict Response to Chemotherapy | Advanced Science | 10.1002/advs.202204097 | 2022 | Mo | Paper | Search Strategy | PDO | 23 | Colorectal Cancer |
| Preclinical In Vivo Validation of the RAD51 Test for Identification of Homologous Recombination-Deficient Tumors and Patient Stratification | Cancer Research | 10.1158/0008-5472.CAN-21-2409 | 2022 | Pellegrino | Paper | Search Strategy | PDX | 9 | Breast Cancer, Pancreatic Cancer |
| A platform for efficient establishment, expansion and drug response profiling of high grade serous ovarian cancer organoids | BioRXiv, Developmental Cell | 10.1101/2022.04.21.489027 | 2022 | Senkowski | Paper | Search Strategy | PDO | 2 | Ovarian Cancer |
| Patient-Derived Xenografts and Organoids Recapitulate Castration-Resistant Prostate Cancer with Sustained Androgen Receptor Signaling | Cells | 10.3390/cells11223632 | 2022 | Van Hemelryk | Paper | Search Strategy | PDX | 2 | Genitourinary Cancer |
| Correlation between drug sensitivity profiles of circulating tumour cell-derived organoids and clinical treatment response in patients with pancreatic ductal adenocarcinoma | European Journal of Cancer | 10.1016/j.ejca.2022.01.030 | 2022 | Wu | Paper | Search Strategy | PDO | 2 | Pancreatic Cancer |
| Application of tumoroids derived from advanced colorectal cancer patients to predict individual response to chemotherapy | Journal of Chemotherapy | 10.1080/1120009X.2022.2045827 | 2022 | Yao | Paper | Search Strategy | PDO | 26 | Colorectal Cancer |
| Proteotranscriptomic analysis of advanced colorectal cancer patient derived organoids for drug sensitivity prediction | Journal of Experimental & Clinical Cancer Research | 10.1186/s13046-022-02591-z | 2023 | Papaccio | Paper | Author Acquired | PDO | 1 | Colorectal Cancer |
| MYC-mediated resistance to trametinib and HCQ in PDAC is overcome by CDK4/6 and lysosomal inhibition | Journal of Experimental Medicine | 10.1084/jem.20221524 | 2023 | Silvis | Paper | Author Acquired | PDX | 1 | Pancreatic Cancer |
| Co-Clinical Study of [fam-] Trastuzumab Deruxtecan (DS8201a) in Patient-Derived Xenograft Models of Uterine Carcinosarcoma and Its Association with Clinical Efficacy | Clinical Cancer Research | 10.1158/1078-0432.CCR-22-3861 | 2023 | Yagishita | Paper | Author Acquired | PDX | 2 | Sarcoma |

**Appendix 2 (Continued) – Information on each included publication.**
